# The social dimension of apathy: Evidence for a distinct domain from 11,243 individuals across health and neurocognitive disorders

**DOI:** 10.1101/2025.09.29.25336885

**Authors:** Sijia Zhao, Rong Ye, Qian-Yuan Tang, Bahaaeddin Attaallah, Sofia Toniolo, Youssuf Saleh, Matthew A. Rouse, Peter Garrard, M. John Broulidakis, Sian Thompson, Sanjay G. Manohar, Sarosh R. Irani, Yuen-Siang Ang, Patricia Lockwood, Matthew A.J. Apps, Panpan Hu, Kai Wang, James B. Rowe, Campbell Le Heron, Masud Husain

## Abstract

Apathy is a highly prevalent and disabling neuropsychiatric syndrome, but its multi-dimensional structure is a challenge for progress towards better identification and treatment. A critical question is whether social disengagement reflects a distinct deficit in social motivation or a by-product of diminished initiative or emotional blunting. Previous studies have been constrained by modest sample sizes and limited use of apathy-specific instruments or phenotypically narrow cohorts. Here, we examined data from 11,243 individuals recruited across multiple centres, including 1,154 neurological patients with Alzheimer’s disease, Parkinson’s disease, frontotemporal dementia, autoimmune encephalitis and small vessel disease, alongside people with depression and healthy adults. Across exploratory and confirmatory factor analyses, symptom-level network modelling, and lifespan analyses, social apathy consistently emerged as a coherent and separable dimension. The pattern was stable across health, depression, and neurocognitive disorders, and evident from adolescence to late life. These findings suggest that social apathy is a distinct and stable component of apathy. Recognising this domain has important implications for diagnostic nosology, for probing the underlying cognitive process that leads to apathy, and its structural correlates, leading ultimately to better targeted psychosocial and pharmacological interventions.

## Introduction

Apathy is a neuropsychiatric syndrome characterised by diminished goal-directed behaviour. It represents one of the most prevalent and debilitating neuropsychiatric features of prodromal and established neurocognitive disorders (NCDs).^1–3^ In older people, its presence can not only be a marker of significant underlying neuropathology but also a key predictor of accelerated functional decline and progression to dementia and death, imposing a profound societal and healthcare burden.^4–9^ There is growing recognition that a spectrum of amotivational states is widely distributed throughout the general population, affecting otherwise healthy adults across the lifespan.^10,11^ Apathy is a distinct clinical entity, dissociated from phenomenologically related conditions such as depression and anhedonia, highlighting the need for a precise characterisation of its core components for targeted therapeutic strategies.^10,12–18^

While most researchers concur that apathy is a multidimensional syndrome, the nature and composition of these dimensions remain a subject of considerable debate.^19^ The conceptualisation of apathy has evolved considerably from its seminal definition by Marin as a “diminished motivation unattributable to emotional distress, cognitive impairment, or diminished consciousness”.^20^ A paradigm shift occurred with the introduction of neurobiologically-grounded models, most notably the framework by Levy and Dubois, which leveraged insights from cognitive neuroscience to separate apathy into subtypes linked with dysfunction in distinct frontostriatal circuits.^21^ This model was instrumental in delineating an emotional-affective dimension from deficits in self-initiated motor activity (which they termed “auto-activation apathy”). It also informed the 2009 diagnostic criteria, which proposed that a diagnosis of apathy required not only a core loss of motivation but also symptoms across at least two of three domains: behaviour, cognition, and emotion.^2^ Subsequently, the 2021 criteria for apathy in NCDs, established via a formal consensus survey of 143 international experts, defined apathy with three clinically heuristic dimensions: diminished initiative (behavioural), diminished interest (cognitive), and diminished emotional expression/responsiveness (emotional).^1^

Social disengagement was not specifically featured in these three dimensions. This omission is striking given that a 2018 transdiagnostic framework for apathy had recognised “Social Interaction” as a distinct aspect of apathy, and a potential fourth dimension in revised criteria.^22^ The decision to exclude “social apathy” from the 2021 criteria was not based on a conceptual rejection, but on a lack of expert consensus regarding its independence.^1^ In the consensus survey, agreement on the question, “Do you agree that [“loss of social activity”] should be considered an independent domain?” was only 59.4%. This figure stands in stark contrast to the consensus for each of the other three dimensions: behavioural (85.7%), cognitive (86.4%), and emotional (94.4%). The workgroup therefore reasoned that “examples of behaviours previously listed under social apathy could also be encompassed by diminished interest, initiative, and emotional expression/responsiveness.”

The decision to omit social apathy as a separate dimension was explicitly provisional. The consensus workgroup agreed that “should evidence arise in the future that demonstrates the need to separate social interactions, that decision would be reconsidered”.^1^ This statement articulates a clear, unresolved, and vital clinical research question: Is social apathy a distinct and separable dimension, or an epiphenomenon of deficits in general initiative and interest, possibly influenced by emotional blunting, applied to a social context? For instance, should “I have lost interest in my friends” be classified under a general loss of interest, or does it tap into a specific deficit in social motivation? This can be tested psychometrically: if social symptoms consistently cluster together and segregate from non-social initiative and interest items, they must be considered a standalone dimension. This validation has not yet been performed with the scale required for a definitive conclusion.^19^

The current study was designed to provide direct, large-scale evidence for the status of social apathy. We first sought to determine the fundamental factor structure of apathy by conducting a detailed, symptom-level analysis of data from three widely used and validated questionnaires—the Apathy Motivation Index (AMI),^23^ the Apathy Evaluation Scale (AES),^24^ and the Dimensional Apathy Scale (DAS)^25^—in a sample of 479 healthy adults. We then leveraged this foundation to investigate the standing of social apathy as a distinct dimension in a large and clinically diverse sample of 11,243 individuals.

Considering the high prevalence of apathy across different clinical populations,^26–28^ we examined this factor structure across three distinct populations: patients with a wide spectrum of NCDs, people without NCD but with depressive symptoms, and healthy individuals spanning the adult lifespan. Our primary hypothesis was that social apathy would emerge as a robust and separable factor across these diverse groups. Such a finding would have significant implications, reframing social apathy from a secondary feature to a core dimension that demands prioritisation in theoretical, experimental, and clinical work.

## Methods

To systematically investigate the dimensional structure of apathy, with a primary focus on determining the validity and stability of a “social apathy” dimension, we proceeded through four sequential steps. We:

1) Conducted an exploratory, data-driven analysis to identify the fundamental dimensions of apathy from a comprehensive set of symptoms in healthy participants
2) Performed a confirmatory analysis to validate the emergent structure within a large clinical cohort
3) Ran a symptom-level network analysis to map the fine-grained architecture of apathy across different populations
4) Conducted a lifespan analysis to assess the stability of this architecture across aging and neurocognitive conditions.

### Participants

This study used a cross-sectional, multi-centre survey design. Ethical approval was granted by the relevant ethics committees for all recruitment sites, including the University of Oxford (IRAS ID: 248379, Ref:18/SC/0448), University of Cambridge (IRAS ID: 252986), University of Birmingham (ERN_20-1897AP16) in UK, Anhui Medical University in China (No. 2022439), and New Zealand Brain Research Institute (HDEC - URB/09/08/037; URB/09/08/037/AM04) in New Zealand. All research was conducted in accordance with the Declaration of Helsinki, and all participants provided written or digital informed consent. This research followed the American Association for Public Opinion Research (AAPOR) Best Practices for Survey Research.

A total of **11,243 participants** were recruited through a combination of online platforms (Prolific), university volunteer databases, and specialist patient clinics. The data were collected between 12 July 2016, and 6 May 2025. This cohort is referred to as the **Full Cohort**, consisted of three sub-groups (see **Table 1** for full demographics):

- **NCDs cohort** (n = 1,154): Patients (mean age 66.3, 23-93^1^) with a range of diagnoses established by experienced neurologists according to consensus criteria. This cohort included Alzheimer’s disease (AD; n=130), Mild Cognitive Impairment (MCI; n=34), Subjective Cognitive Decline (SCD; n=136) ^2^, Parkinson’s disease (PD; n=551), Dementia with Lewy Bodies (DLB; n=39), Frontotemporal Dementia (FTD; n=89), Autoimmune Encephalitis (AIE; n=86), and Small Vessel Disease (SVD; n=89). The majority of neurological patients were recruited from the Cognitive Disorders Clinic at John Radcliffe Hospital, Oxford, UK. All AIE patients (N = 86) were recruited from Autoimmune Neurology Clinic, John Radcliffe Hospital, Oxford, UK.^29,30^ Forty-seven FTD patients were recruited from specialist dementia clinics in Addenbrooke’s Hospital, Cambridge, UK and St George’s Hospital, London, UK (N = 4). Additionally, 195 PD patients were recruited from the New Zealand Parkinson’s Progression Programme (NZP3) in Canterbury, New Zealand,^31^ and 192 PD patients were recruited from the China Parkinson’s Disease Advanced Center in the First Affiliated Hospital of Anhui Medical University, Anhui, China.
- **Depression Cohort** (Depression; n = 1,324): Individuals (mean age 22.3, 16-81) who either had a formal diagnosis of Major Depressive Disorder (MDD), confirmed by two independent psychiatrists according to DSM-IV criteria, or who scored above established cut-offs on depression screening tools.
- **Healthy Control Cohort** (HC; n = 8,765): Community-dwelling individuals (mean age 39.3, 16-93) self-reported without major neurological or psychiatric disorders.

**Table 1:** Demographics and average measures for the two studies in the present study. For continuous variables (e.g., age, years of education, questionnaire scores), the mean and standard deviation (SD) are reported, along with the number of participants for whom data were available (indicated in parentheses). Gender distribution is reported as the number of female (F) and male (M) participants, non-binary (NB) participants. Abbreviations: N.A. = Data missing. AMI = Apathy-Motivation Index, ACE = Addenbrooke Cognitive Examination-III.

| Study | Group | N | Age | Gender | AMI Total | ACE Total |
| --- | --- | --- | --- | --- | --- | --- |
| **Study 1** | **HC** | **479** | **29.7 (18-74)** | **F:249, M:230, NB:0** | **1.4 (0.2-3.2)** |  |
| **Study 2** | **(Below)** | **11243** | **40.1 (16-93)** | **F:5417, M:4948, NB:17** | **1.4 (0.0-4.0)** | **84.3 (2.0-100.0) [N=739]** |
|  | HC | 8765 | 39.3 (16-91) | F:4251, M:3740, NB:16 | 1.4 (0.0-3.8) | 96.9 (88.0-100.0) [N=121] |
|  | Depression | 1324 | 22.3 (16-81) | F:745, M:578, NB:0 | 1.5 (0.2-4.0) | 91.0 (49.0-100.0) [N=12] |
|  | Neuro. | 1154 | 66.3 (23-93) | F:421, M:630, NB:1 | 1.6 (0.1-3.8) | 81.6 (2.0-100.0) [N=606] |
|  | AD | 130 | 69.7 (49-92) | F:66, M:63, NB:1 | 1.5 (0.1-3.4) | 69.2 (16.0-97.0) [N=84] |
|  | MCI | 34 | 67.6 (54-90) | F:18, M:16, NB:0 | 1.5 (0.9-3.1) | 84.7 (75.0-95.0) [N=25] |
|  | SCD | 136 | 57.6 (23-78) | F:69, M:65, NB:0 | 1.6 (0.4-3.0) | 91.8 (63.0-100.0) [N=104] |
|  | PD | 551 | 67.5 (39-88) | F:169, M:286, NB:0 | 1.8 (0.2-3.8) | 86.8 (20.0-100.0) [N=147] |
|  | DLB | 39 | 68.8 (57-93) | F:10, M:28, NB:0 | 1.5 (0.3-2.7) | 77.0 (26.0-93.0) [N=25] |
|  | FTD | 89 | 63.9 (42-90) | F:31, M:56, NB:0 | 1.4 (0.1-3.5) | 59.7 (2.0-98.0) [N=76] |
|  | AIE | 86 | 63.1 (26-86) | F:23, M:62, NB:0 | 1.4 (0.4-2.8) | 91.2 (75.0-100.0) [N=70] |
|  | SVD | 89 | 70.5 (52-92) | F:35, M:54, NB:0 | 1.4 (0.3-2.8) | 85.1 (25.0-100.0) [N=75] |

### Measurements

#### Assessment of apathy

For the primary, cross-sectional analysis of the full cohort (*N* = 11,243), we used the Apathy Motivation Index (AMI) as a self-report measure of apathy.^23^ The AMI is an 18-item instrument designed to assess three *a priori* domains of apathy: *Behavioural* (e.g., “When I decide to do something, I am able to make an effort easily”), *Social* (e.g., “I start conversations without being prompted”), and *Emotional* (e.g., “I feel bad when I hear an acquaintance has an accident or illness”), noting that each question in isolation may reflect a combination of cognitive and behavioural elements.

To facilitate a more comprehensive phenotyping of apathy and to establish convergent validity, a subsample of 479 healthy participants also completed two additional, widely used apathy scales: the Apathy Evaluation Scale (AES)^24^ and the Dimensional Apathy Scale (DAS).^32^

The selection of these specific instruments was theoretically motivated: (1) AES, an 18-item self-report questionnaire, was grounded in the foundational conceptualisation of apathy proposed by Marin, and designed to assess the behavioural, cognitive and emotional dimensions of the syndrome.^20,24^ (2) DAS, a 24-item instrument, was based on the influential, neurologically informed framework proposed by Levy and Dubois and dissociated apathy into auto-activation apathy (impaired self-initiation of thoughts or actions), executive apathy (impaired planning and organisation), emotional-affective apathy (affective indifference).^21,32^

This multi-instrument approach, leveraging the unique theoretical underpinnings of AMI, AES and DAS was implemented to ensure a comprehensive and multidimensional characterisation of apathy.

#### Cognitive Function

A subset of NCD patients (N=739) completed the Addenbrooke’s Cognitive Examination-III (ACE-III), a 100-point scale of overall cognition, with subscales assessing attention, memory, verbal fluency, language, and visuospatial skills.^33^

### Statistical Analysis

Statistical analyses were conducted with MATLAB (R2025a) and R (version 4.5.1).

#### Exploratory factor analysis

To investigate the latent structure of apathy symptoms, an EFA was performed on the **Healthy Control Cohort** who completed all three apathy questionnaires: AMI, AES, and DAS (N=479). Suitability for factor analysis was confirmed using the Kaiser-Meyer-Olkin (KMO) measure and Bartlett’s Test of Sphericity. The number of factors to retain was determined by Kaiser’s Criterion (Eigenvalues > 1). Given the theoretical expectation that apathy dimensions would be related, an oblique (promax) rotation was applied to the factor structure to allow for correlations between the latent factors.

#### Confirmatory factor analysis (CFA)

This was performed on the AMI data from **NCDs cohort**, using the R lavaan package. We specified an *a priori* three-factor model based on previous findings,^26^ composed of Behavioural, Social, and Emotional factors. Unlike the exploratory analysis, which combined items from AMI, AES, and DAS to test the broad latent structure of apathy, the CFA was restricted to AMI data; because the AMI only contains items reflecting these three domains, the confirmatory model was designed specifically to test this structure rather than to reproduce the five-factor solution from the exploratory stage. Three items (AMI-2, AMI-6, AMI-8) showed lower indicator reliability within their designated factors, with weak or inconsistent loadings and elevated residual correlations. Following standard practice, these items were removed to improve model fit and ensure that each latent construct was represented by items with high reliability and factorial coherence. To account for potential non-normality in the data, we used a robust Maximum Likelihood estimator, which provides scaled test statistics and robust standard errors. Model adequacy was assessed using a standard battery of goodness-of-fit indices: the robust chi-square (χ2) test, the Comparative Fit Index (CFI), the Tucker-Lewis Index (TLI), the Root Mean Square Error of Approximation (RMSEA) with its 90% confidence interval, and the Standardized Root Mean Square Residual (SRMR). Conventional criteria were used to evaluate model fit, with CFI/TLI values ≥ 0.90, RMSEA ≤ 0.08, and SRMR ≤ 0.06 considered indicative of acceptable fit. Standardised factor loadings (λ) and inter-factor correlations were examined to assess the coherence and distinctiveness of the latent constructs.

#### Factor purity

To quantify the specificity of items to their designated latent construct, we calculated a factor purity index for each item, defined as the squared loading of an item on its primary designated factor, divided by its communality (the sum of its squared loadings across all factors). A higher purity index, approaching 1.0, indicates that an item’s explained variance is almost exclusively attributable to a single factor. The mean factor purity and its standard error of the mean (SEM) were then calculated for each factor to compare their relative factorial purity. An initial one-way ANOVA was conducted to test for any significant differences in mean purity scores across the factors. Following this, a planned comparison was performed using a Welch’s independent samples t-test. This test specifically evaluated whether the mean purity of the Social Apathy factor was significantly higher than the combined mean of the remaining four factors. The alpha level for determining statistical significance was set at 0.05 for all tests.

#### Network estimation and visualisation

To investigate the symptom-level structure of apathy in the **Full Cohort** (N=11,243), we constructed weighted, undirected networks from the 18 AMI items for the HC, Depression and NCD groups separately. Nodes represented items, and edge were weighted by the absolute Spearman correlation coefficients, which is robust to non-normal data distributions typical of Likert scales. For visualisation, a sparse network was generated by retaining only edges with a correlation of ∣ρ∣≥0.1. The network was visualised in R using the qgraph package with the Fruchterman-Reingold algorithm, a force-directed layout that positions strongly connected nodes closer together. Nodes were coloured based on their a priori AMI domain (Behavioural, Social, Emotional). Node size was scaled to represent its **between-domain connectivity**, defined as the mean absolute Spearman correlation of that node with all nodes from the other two domains, visually highlighting bridging items.

#### Module and centrality detection

To empirically identify functional clusters within the network, we applied a Louvain-style modularity maximisation algorithm with local greedy optimisation in a single iterative pass. This data-driven community detection algorithm iteratively partitions the network to maximise modularity, a metric that quantifies the density of intra-community connections relative to inter-community connections. This aligns with the conceptual goal of identifying symptom clusters that co-occur more strongly with each other than with symptoms outside the cluster. Modularity (*Q*) is formally defined as:

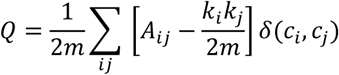

where *A_ij_* represents the weight of the edge between nodes *i* and *j*, *k_i_* is the sum of weights of all edges connected to nodes *i* (strength), *m* is the sum of all edge weights in the network, and *δ*(*c_i_*, *c_i_* ) is 1 if nodes *i* and *j* belong to the same community (*c*_*i*_ = *c*_*i*_ ) and 0 otherwise. Within each module, we defined the central node as the one with the highest intra-modular strength, i.e., the sum of its connection weights to all other nodes in the same community.

#### Lifespan trajectory analysis

To assess the stability of the apathy network structure across age, we employed a continuous, age-based sliding-window analysis on the AMI data for each of the three cohorts. The analysis systematically progressed across the age range of each group in 1-year increments, using a 10-year window weighted by a Gaussian kernel (σ = 5 years). Within each age window, we re-computed the network. To ensure a robust result, the Louvain algorithm was run 1000 times, and the most frequently detected module partition was selected for all subsequent analyses. This stable partition was used to determine the number and composition of modules for that specific age. Any age window with a weighted effective sample size below 30 was excluded from the analysis to ensure statistical stability.

## Results

### Establishing the fundamental structure of apathy with exploratory factor analysis

We first sought to derive the fundamental phenomenological structure of apathy symptoms, unconstrained by the theoretical assumptions of any single assessment scale. Exploratory Factor Analysis (EFA) was conducted on **the Healthy Control Cohort** (N=479) on the combined 60 items from three widely used apathy scales: the AES, DAS, and AMI.

This multi-instrument approach was a strategic choice. The AES was grounded in Marin’s foundational tripartite model (behavioural, cognitive, emotional),^20,24^ while the DAS was based on the influential neurobiologically-informed framework of Levy and Dubois, which divided apathy into auto-activation, executive, and emotional-affective subtypes.^21,32^ The AMI is the only one of these scales that explicitly includes a named social domain, although social apathy is partially and indirectly represented in questions of the other scales.^19^ For example, the AES and DAS contain items with social relevance (e.g., AES: “Getting together with friends is important to me”; DAS: “I contact my friends”), albeit with limited coverage of social initiation, maintenance of social ties, and social motivation. By combining all 60 items, we created a rich symptom catalogue to comprehensively characterise apathy and formally test a core hypothesis.

Specifically, if social apathy were merely a facet of behavioural or emotional apathy, its corresponding items could load onto those factors. For instance, diminished initiative in conversation (“I start conversations without being prompted”) would be absorbed into a general behavioural/initiative factor, while reduced enjoyment from social interaction (“I enjoy doing things with people I have just met”) would be subsumed by an emotional or motivational anhedonia factor. The emergence of a distinct, statistically independent social factor would, therefore, provide evidence for its status as a separate dimension. Alternatively, a separate factor could arise for methodological reasons—for example, if socially oriented items differed systematically in style, linguistic complexity, or word frequency. Such possibilities can be examined post hoc if an additional factor is identified.

The data were well-suited for factor analysis, as confirmed by a high KMO measure of sampling adequacy (KMO = 0.90) and a significant Bartlett’s Test of Sphericity (χ²(1770) = 12640.7, p < .001). The EFA revealed a clear and interpretable five-factor structure that explained the data well (χ² = 3411.6, df = 1480, p < 0.001). These five dimensions were interpreted as reflecting:

1. Behavioural Apathy: Diminished self-initiated, goal-directed behaviour.
2. Cognitive Apathy: Diminish expression of interest, “wanting”, and curiosity.
3. Social Apathy: Diminished social interaction and engagement.
4. Emotional Apathy: Emotional blunting and affective indifference.
5. Executive functions: Difficulties with concentration, planning, and organisation.

As shown in **Figure 1**, items assessing social engagement loaded strongly and cleanly onto a single, distinct Social Apathy factor. For example, AMI Question 14 (“I start conversations without being prompted”) and AES Question 13 (“Getting together with friends is important to me”) anchored this factor. This social dimension was clearly separable from the Behavioural dimension, which included items such as AMI Question 15 (“When I have something I need to do, I do it straightaway…”) and DAS Question 18 (“I keep myself busy”). Social Apathy was also distinct from the Cognitive dimension, defined by items such as AES Question 9 (“I spend time doing things that interest me.”) and AES Question 5 (“I am interested in learning new things”). A complete list of item loadings is provided in **Supplementary Table 1**.

**Figure 1.**
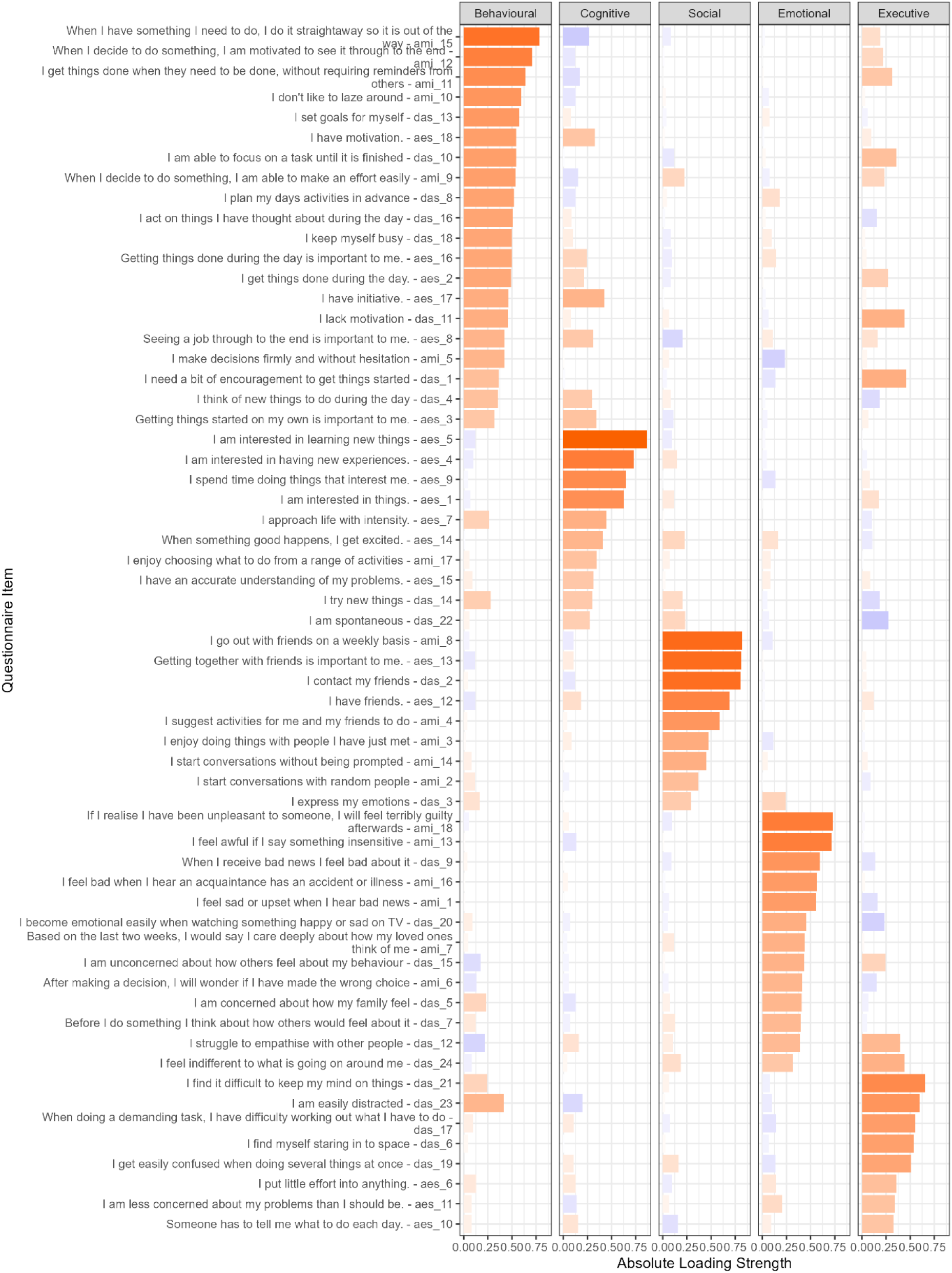
Factor structure of apathy items. Results from an exploratory factor analysis with varimax rotation on all items from the Apathy Motivation Index (AMI), Dimensional Apathy Scale (DAS), and Apathy Evaluation Scale (AES) in 500 healthy adults. Positive factor loadings are shown in orange and negative loadings are shown in purple. The order of the items was arbitrary. The loading strength for each item can be found in Supplementary Table 1.

Analysis of the inter-factor correlations revealed a nuanced pattern of relationships (**Table 2**). Using thresholds of ≥0.3 for moderate and ≥0.5 for strong relationships,^34^ we observed strong correlations among Behavioural, Cognitive, and Social apathy (r = 0.49 to 0.59). This may suggests a higher order “Initiative/Auto-activation” cluster like proposed by Levy and Dubois.^21^ In contrast, Emotional and Executive apathy showed weak correlations with all other factors (all |r| < 0.18), and Executive apathy even had a slight negative relationship with Social apathy (r = -0.11), underscoring their dissociation from the other domains.

**Table 2:** Inter-factor correlation matrix. Correlation coefficients for the five factors identified through EFA. Values in bold indicate moderate (r ≥ 0.3) to strong (r ≥ 0.5) relationships between the apathy dimensions.

|  | Factor1<br>Behavioural | Factor2<br>Cognitive | Factor3<br>Social | Factor4<br>Emotional | Factor5<br>Executive |
| --- | --- | --- | --- | --- | --- |
| Factor1 Behavioural apathy | 1 | **0.589** | **0.494** | 0.119 | 0.1674 |
| Factor2 Cognitive apathy | **0.589** | 1 | **0.579** | 0.1751 | 0.1235 |
| Factor3 Social apathy | **0.494** | **0.579** | 1 | 0.299 | -0.1074 |
| Factor4 Emotional apathy | 0.119 | 0.175 | 0.299 | 1 | 0.0506 |
| Factor5 Executive apathy | 0.167 | 0.123 | -0.107 | 0.0506 | 1 |

To quantify the robustness of this factor structure, we analysed factor purity—a measure of how specifically items relate to their primary dimension. An initial one-way ANOVA showed no overall difference in purity scores across the five factors (F(4, 55) = 1.44, p = 0.23). However, post-hoc testing revealed that the mean purity of the Social Apathy factor (M = 88.3%, SEM = 5.0%) was significantly higher than the overall mean of the other four factors (M = 73.8%; t(13.1) = 2.56, p = 0.012), indicating its high factorial coherence in this sample (**Figure 2**).

**Figure 2:**
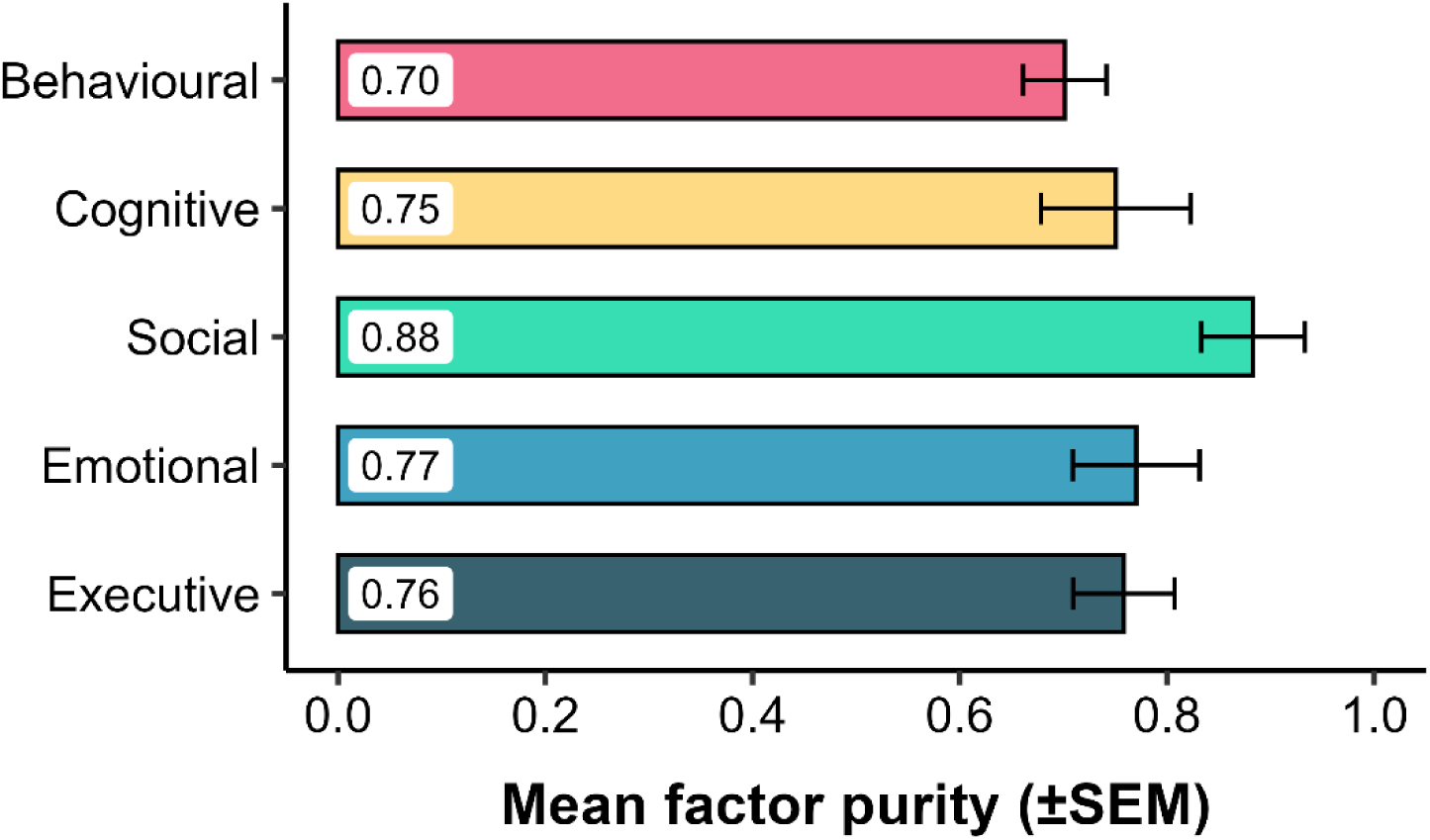
Coherence of the five apathy dimensions as measured by factor purity. A comparison of the mean factor purity scores for the five apathy factors identified in EFA. Factor purity is an index that quantifies the proportion of an item’s shared variance (i.e., its communality) that is attributable to its primary factor. A score approaching 1.0 indicates that an item loads almost exclusively on a single factor, reflecting high dimensional coherence. Each bar represents the mean purity score, averaged across all items constituting that factor, with error bars denoting the standard error of the mean (SEM). The results highlight the significantly higher factorial coherence of the Social Apathy dimension compared to the others. For a detailed breakdown of the factor loadings for each item, see **Supplementary Figure 1**.

Overall, this comprehensive exploratory analysis provides strong evidence that social apathy is not an artefact of a specific questionnaire but emerges as a distinct, coherent, and robust dimension when a wide range of apathy symptoms are considered, and is fundamentally dissociable from behavioural and emotional dimensions.

### Validating the social apathy domain in NCD with confirmatory factor analysis

Having established the existence of a distinct social apathy dimension in a healthy sample, we next sought to determine if this structure holds in a large, clinically relevant population. Not all participants completed multiple apathy questionnaires; in patients, time constraints limited assessment to a single scale. We therefore focused on the AMI, which uniquely includes dedicated social items and has prior evidence supporting a three-factor structure (Behavioural, Social, Emotional) in NCDs.^26,35^ Based on this foundation, we performed a stringent Confirmatory Factor Analysis (CFA) on this a priori model in our large NCD sample (N = 1,154). This analysis was not intended to reproduce the broader five-factor solution from the exploratory stage, since the AMI does not include dedicated items for cognitive or executive apathy, but rather to test whether the three domains represented within the AMI form distinct and coherent latent constructs in patients.

The 3-factor model demonstrated an acceptable to good fit to the data, as indicated by a battery of standard goodness-of-fit indices (CFI = 0.915; SRMR = 0.060; TLI = 0.90; RMSEA = 0.075 [90% CI, 0.070-0.081]). Although the model’s chi-square test was significant (χ²(87) = 624.22, p < .001), this is an expected outcome in very large samples and does not invalidate the model when other fit indices are strong.

Importantly, all individual items demonstrated substantial and statistically significant standardised loadings on their intended latent factors—Behavioural (λ = 0.43-0.82), Emotional (λ = 0.59-0.85), and Social (λ = 0.42-0.81). This confirms the high internal coherence of each subscale. The analysis also revealed moderate-to-high inter-factor correlations (r = 0.64 to 0.77), indicating that while the three dimensions partially capture distinct aspects of the syndrome, they are not independent.

These CFA results provide strong psychometric validation for the AMI’s three-factor model, confirming that social apathy stands as a distinct and measurable construct within the complex syndrome of apathy in NCDs.

### Mapping the symptom-level architecture across populations

Factor analysis models symptoms as indicators of an unobserved latent variable. To gain a more nuanced, bottom-up understanding of the relationships *between* individual symptoms, we turned to network analysis.^36,37^ This approach models apathy as a complex system of interacting symptoms, providing a fine-grained map of symptom-to-symptom associations. This perspective has been shown to capture clinical complexity more directly, explain comorbidity through bridge symptoms, and identify central targets for intervention.^36^ We therefore asked whether the dimensional structure identified via CFA would be replicated at the symptom-network level, providing convergent evidence for the distinct role of social apathy.

A network analysis on the AMI items in our entire sample (N=11,243) stratified into three major cohorts: NCD, Depression and HC (**Figure 3**). We used community detection algorithms to identify “modules”—clusters of symptoms that are more strongly correlated with each other than with other symptoms in the network. This is conceptually analogous to identifying factors but is derived directly from the network structure without assuming a latent cause.

**Figure 3:**
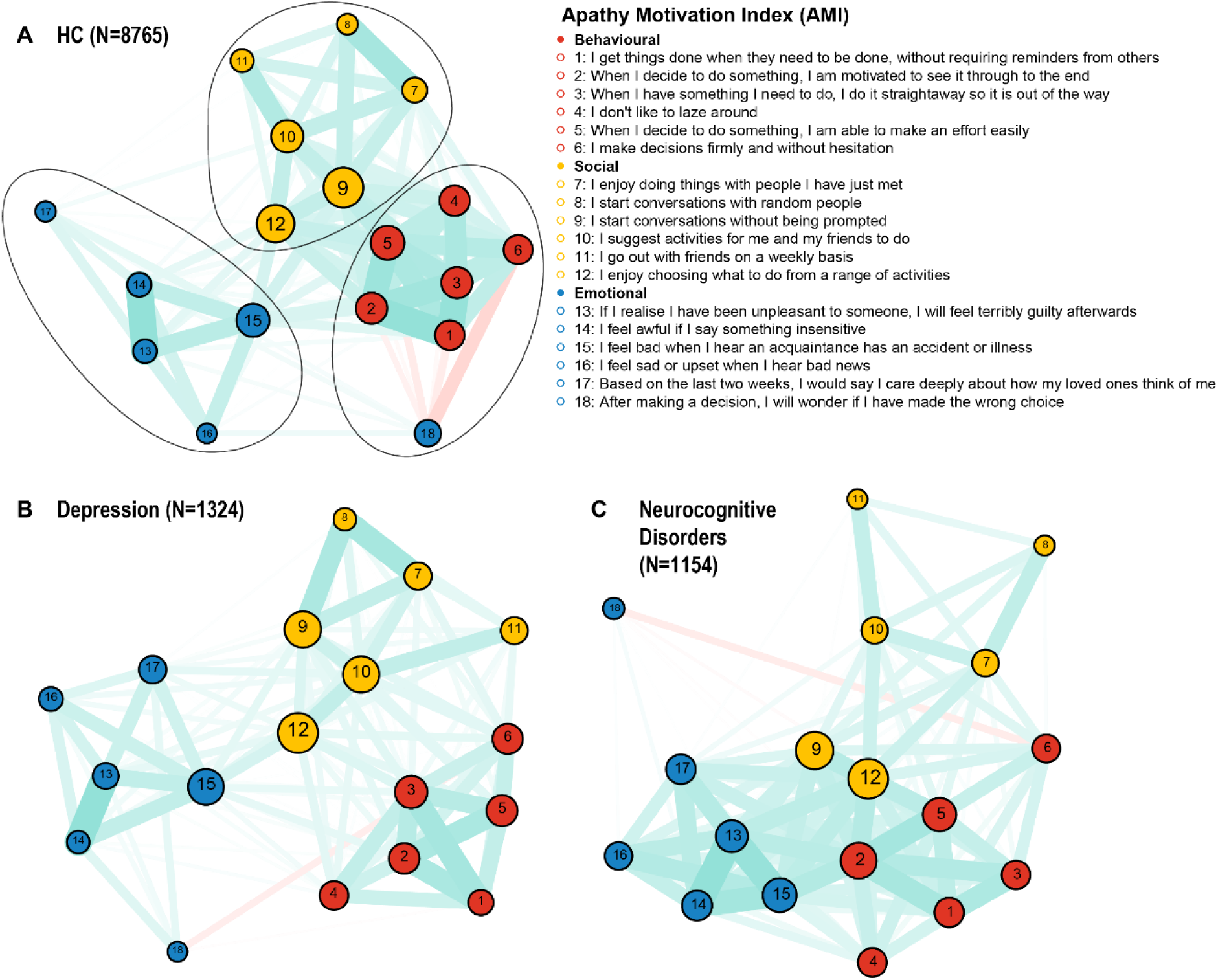
Apathy symptom networks of the AMI across three cohorts. Network structures of Apathy Motivation Index (AMI) items for (A) Healthy Controls, (B) Depression, and (C) Neurocognitive Disorders. Nodes (circles) represent individual symptoms and are coloured by their a priori domain: red=Behavioural, yellow=Social, blue=Emotional. Edges (lines) represent Spearman correlations between symptoms, thresholded for clarity at a strength of ∣ρ∣≥0.1. Cyan edges denote positive correlations, while red edges denote negative ones. Edge thickness is proportional to the correlation strength. The layout places strongly related symptoms closer together, visually highlighting the distinct clustering of the three apathy domains.

To ensure the robustness of our findings, we applied the Louvain community detection algorithm with 1000 random restarts to the item correlation network of each cohort.^38,39^ This stability analysis revealed robust and distinct module structures for each group. The NCD cohort’s network was exceptionally stable, identifying a three-module solution in 100% of iterations, which corresponded precisely to the a priori Behavioural, Social, and Emotional dimensions of the AMI. The Depression cohort most consistently revealed a four-module solution (80.9% of runs), which often separated the behavioural items into sub-groups but consistently preserved the integrity of the Social and Emotional modules. In the HC cohort, a three-module solution was most frequent, found in 70.0% of iterations, again mapping cleanly onto the Behavioural, Social, and Emotional domains.

### The stability of the social apathy module across the lifespan

A final question was whether the dissociation of social apathy is a stable feature or if it varies with age. For example, do social symptoms merge with emotional or behavioural domains during adolescence or in late life? Using a sliding-window analysis across the lifespan (ages 16 to 90), we constructed separate apathy network for overlapping age windows in each of the three major cohorts. Within each age-specific network, we performed community detection to determine if a distinct “Social Apathy” module was present and to quantify its coherence.

The results, as shown in **Figure 4**, show that the Social Apathy dimension was identifiable at every age point across all three cohorts. This module had a social item as its central (most connected) node and was consistently characterised by high domain purity (the proportion of social items within the module). This provides convergent visual evidence that the segregation of social apathy symptoms is not confined to a particular life stage, but emerges consistently in the symptom network across adolescence, adulthood, and late life.

**Figure 4:**
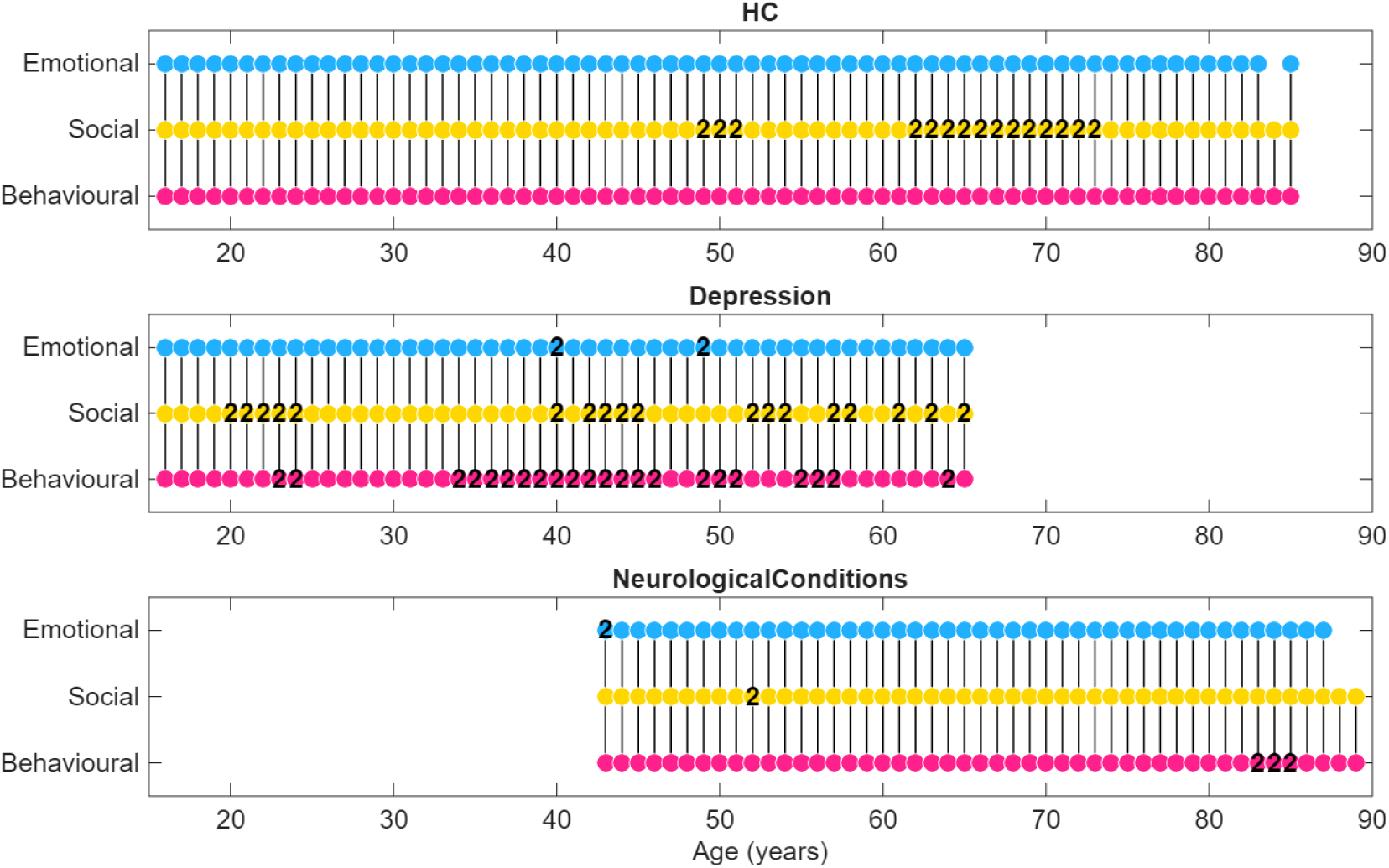
Stability of social apathy across the lifespan. Domains of apathy modules detected via a sliding-window network analysis across three cohorts: (A) Healthy Controls, (B) Depression, and (C) Neurocognitive Disorders. Each point on the x-axis represents the centre of a 10-year age window for which a symptom network was constructed. Dots are colour-coded by the dominant domain of the module: Behavioural (pink), Social (yellow), or Emotional (blue). Numbers within dots indicate how many distinct modules of that domain were detected in that age window (e.g., “2” means two independent modules of the same domain were present). Critically, if social apathy symptoms had blended into other domains, this would be reflected as breaks or gaps in the social (yellow) row, meaning no independent social module was detected for that age or cohort. The uninterrupted presence of yellow dots across all ages and groups shows that social apathy consistently emerged as its own symptom-centred module, demonstrating its stability as a distinct dimension throughout the lifespan.

To formally test the factors influencing module coherence, we performed a two-way ANOVA with Module Purity as the dependent variable and Group (NCD, Depression, HC) and Domain (Behavioural, Social, Emotional) as factors. The analysis revealed a significant main effect of Domain (F(2, 551) = 68.15, p < .001), as Social and Emotional modules consistently demonstrated higher purity than Behavioural modules. We also found a significant main effect of Group (F(2, 551) = 15.26, p < .001), with module purity being highest in healthy controls and lower in the clinical groups. Finally, a significant Group x Domain interaction (F(4, 551) = 2.56, p = .038) indicated that the precise relationship between domain purity and clinical status was not uniform across the cohorts (see **Supplementary Tables 2-4** for detailed values).

The social module’s purity remained high across all groups (average purity = 0.97 in HCs, 0.94 in Depression, and 0.91 in NCD). This underscores our primary observation: the tendency for social apathy symptoms to cluster together is a stable and fundamental feature of human apathy, present across health and disease and persistent through the adult lifespan.

## Discussion

This study provides large-scale evidence that social apathy is a distinct and cohesive dimension of the apathy syndrome. Across a clinically diverse cohort of over 11,000 individuals, social apathy emerged as a psychometrically robust construct, separable from behavioural and emotional domains. This was observed across the adult lifespan and was preserved in healthy participants, participants with depression, and in participants with a wide range of neurocognitive disorders. Our findings demonstrate that while social apathy is correlated with behavioural apathy, it does not merge with it. The high factorial purity of the social apathy dimension underscores its internal coherence and validates its status as a standalone dimension.

This psychometric distinction is also consistent with emerging neurobiological evidence. General apathy has consistently been associated with dysfunction in a core network for effort-based decision-making for self, centred the dorsal anterior cingulate cortex, ventral striatum, and related frontosubcortical structures.^40,41^ By contrast, converging work highlights a key role for anterior cingulate gyrus (ACCg) in motivating prosocial effort. Neurophysiology and fMRI studies show that ACCg encodes the costs and values of *others’* rewards and effort, but not one’s own.^42,43^ Consistent with this, reduced willingness to exert effort for others correlates with higher social apathy scores on the AMI, the same instrument used in our analyses.^44^ Lesion studies further implicate the ventromedial prefrontal cortex (vmPFC), where damage leads to diminished prosocial motivation—patients earn less for others, discount others’ rewards more steeply, and exert less force when helping, with medial and lateral subregions exerting opposing influences.^45^ Taken together, these findings nominate a distributed socio-motivational circuit, spanning ACCg and vmPFC, as a candidate substrate for the selective loss of social motivation in apathy. Testing this directly through multimodal imaging, lesion models, and longitudinal designs will be essential. At the same time, this motivational system should be distinguished from the broader “social brain” network—including the vmPFC, amygdala, posterior superior temporal sulcus, temporal pole, and temporoparietal junction—which is more typically involved in representing others’ mental states and social knowledge. ^46–48,42^ Although pathology in this socio-affective network can alter social cognition and behaviour,^45,49,50^ the evidence reviewed above points specifically to ACCg and vmPFC as critical for social motivation.

In addition, this dimension’s clinical utility is highlighted by its high agreement between patient and caregiver reports.^26^ This concordance persisted even in conditions like behavioural-variant frontotemporal dementia (bvFTD), where changes in insight can create large discrepancies in the responses to questionnaires about behavioural change.^26^

Our data-driven analysis across three apathy scales (AMI, AES, and DAS), where that data was available in our cohorts, resolved a clear five-factor structure: behavioural, cognitive, social, emotional apathy and executive impairments. This is consistent with a more granular organisation of apathy that refines, rather than contradicts, the influential three-part framework of Levy and Dubois.^21^ The strong correlations among the behavioural, cognitive, and social domains (r = 0.49–0.59) may represent distinct expressions of a broader “auto-activation” or “initiative” domain. Under this view, social apathy reflects a unique manifestation of impaired initiative in the social domain, but its coherence and factorial purity indicate that it cannot be reduced to behavioural or cognitive apathy alone. Future work could formally test this hierarchical structure using higher-order factor models, which may help reconcile the separability of social apathy with its overlaps with general motivational deficits. Clinically, this implies that while treatments targeting general initiative (e.g. dopaminergic agents or behavioural activation) may improve all three domains, additional social-specific interventions may be required to address the unique variance captured by social apathy.

However, several limitations must be acknowledged. First, the study was cross-sectional and relied on symptom survey data, which precludes causal inferences about the development of apathy and is subject to the inherent biases of self-report. Some disorders lead to violations of the assumptions underlying questionnaire responses.^51^ Symptoms constitute the key features of a neuropsychiatric diagnosis such as apathy and this remains but further work is necessary to establish if each symptom domain maps selectively to different brain circuits. Second, while psychometrically robust, the AMI does not offer the same granular assessment of cognitive apathy as scales like the AES and executive apathy like DAS. Nevertheless, the five-factor structure that emerged in this analysis shows that none of the scales on their own covers all five domains comprehensively. Third, differences in the questions related to social apathy may introduce confounding factor structures, if for example they were to differ in grammar, frequency or complexity of the words used in items. This risk is mitigated by the ubiquity of the social apathy factor in health and disease states.

Recognising social apathy as a distinct domain also has important practical implications. From an assessment perspective, diagnostic protocols could be expanded to include dedicated measures of social apathy, allowing clinicians to differentiate it from superficially similar conditions such as anhedonia (reduced pleasure from social interaction), social anxiety (avoidance due to fear rather than lack of motivation), or depression (where social withdrawal is a core symptom). Clinically, this may enable more accurate prognosis: social disengagement could prove a stronger predictor of loneliness, caregiver burden, and quality-of-life decline than other apathy domains. In turn, recognising social apathy could guide targeted interventions. While treatments enhancing general initiative (e.g. dopaminergic agents or behavioural activation) may benefit multiple domains, interventions such as structured group-based programmes (e.g. psychosocial engagement game), caregiver-mediated engagement, or digital tools (e.g. social robots, telepresence technologies) may be suited to ameliorating social apathy. Finally, including a social dimension in diagnostic criteria would ensure systematic measurement across clinical trials, stimulate research into the specific brain circuits underlying social motivation, and refine phenotypic descriptions of disorders in which social changes are early and prominent, such as frontotemporal dementia and Parkinson’s disease.

In conclusion, we propose that social apathy is a distinct and stable dimension of the apathy syndrome, not reducible simply to diminished initiative, interest, or emotional responsiveness. These findings provide a strong mandate for reinstating a social dimension into the diagnostic framework for apathy in NCDs. Recognising this dimension is a critical step toward a more precise understanding of its unique neurobiology and, most importantly, opens the door for the development of targeted psychosocial and pharmacological interventions to address the profound loss of social connection that defines this debilitating condition.

## Data Availability

The analysis code and data from healthy participants that support the conclusions of this article will be openly available via the Open Science Framework (OSF) upon publication. Patient data are available upon reasonable request. Access is contingent upon agreement with the original data providers (i.e. the researchers responsible for data collection) and subject to applicable ethical, legal, and confidentiality constraints.

## Acknowledgements

This work was supported by the Wellcome Trust and the National Institute for Health Research (NIHR) Oxford Health Biomedical Research Centre. S.Z., S.T., M.J.B., and M.H. are funded by the Wellcome Trust [226645/Z/22/Z]. S.T., S.G.M. and M.H. are funded by and the NIHR Oxford Health BRC. S.G.M. is also supported by the NIHR Oxford BRC. J.B.R. is supported by the Medical Research Council [MC_UU_00030/14; SUAG/092 G116768], the Wellcome Trust [220258], the NIHR Cambridge Biomedical Research Centre [NIHR203312], and the Holt Fellowship. S.R.I. is supported by a Senior Clinical Fellowship from the Medical Research Council [MR/V007173/1], a Wellcome Trust Fellowship [104079/Z/14/Z], the Kogod Centre on Aging (Mayo Clinic), and the NIHR Oxford Biomedical Research Centre. R.Y. is supported by the National Natural Science Foundation of China [82171917, 82471271, U23A20424], the Anhui Provincial Natural Science Foundation [2408085Y047], and the Natural Science Research Project of Anhui Educational Committee [2023AH050592]. Q.Y.T. was funded by the Guangdong–Hong Kong Universities “1+1+1” Joint Funding Programme. C.L.H. is supported by a grant from the Canterbury Medical Research Foundation [02/2019]. B.A. is supported by an NIHR Academic Clinical Lectureship.

We gratefully acknowledge all the participants, their families, and the staff at the Cognitive Disorders Clinic and the Autoimmune Neurology Clinic at John Radcliffe Hospital, Addenbrooke’s Hospital, St George’s Hospital, the New Zealand Brain Research Institute and the China Parkinson’s Disease Advanced Center for their dedication to this programme.

The views expressed are those of the author(s) and not necessarily those of the NHS, the NIHR, or the Department of Health and Social Care.

## Supplementary Materials

**Supplementary Table 1:** Factor loading of all AMI, AES and DAS items.

| VariableName | Measure | Item | Factor1 | Factor2 | Factor3 | Factor4 | Factor5 |
| --- | --- | --- | --- | --- | --- | --- | --- |
| ami_3 | AMI | I enjoy doing things with people I have just met | 0.022615 | 0.087059 | 0.470036 | -0.11709 | -0.03044 |
| ami_8 | AMI | I go out with friends on a weekly basis | -0.06006 | -0.1043 | 0.822674 | -0.10704 | 0.005834 |
| aes_1 | AES | I am interested in things. | -0.06846 | 0.626662 | 0.117052 | 0.005072 | 0.175286 |
| aes_4 | AES | I am interested in having new experiences. | -0.09798 | 0.730458 | 0.144628 | -0.04469 | -0.05254 |
| aes_5 | AES | I am interested in learning new things | -0.12055 | 0.867387 | -0.09428 | -0.02962 | -0.01348 |
| aes_12 | AES | I have friends. | -0.12127 | 0.183978 | 0.693854 | -0.02724 | 0.124381 |
| aes_13 | AES | Getting together with friends is important to me. | -0.11804 | 0.107727 | 0.814611 | 0.015187 | 0.045554 |
| das_5 | DAS | I am concerned about how my family feel | 0.235757 | -0.13308 | 0.075696 | 0.409869 | -0.06864 |
| das_14 | DAS | I try new things | 0.279572 | 0.303325 | 0.20428 | -0.05169 | -0.18713 |
| das_24 | DAS | I feel indifferent to what is going on around me | -0.08317 | 0.041625 | 0.186009 | 0.316871 | 0.439922 |
| ami_4 | AMI | I suggest activities for me and my friends to do | 0.038629 | 0.044798 | 0.590646 | -0.00035 | 0.03078 |
| das_4 | DAS | I think of new things to do during the day | 0.352652 | 0.297424 | 0.078201 | -0.03231 | -0.18176 |
| ami_5 | AMI | I make decisions firmly and without hesitation | 0.419111 | 0.013753 | 0.063845 | -0.23427 | 0.052522 |
| ami_17 | AMI | I enjoy choosing what to do from a range of activities | 0.062616 | 0.347355 | 0.072575 | 0.086285 | -0.015 |
| aes_7 | AES | I approach life with intensity. | 0.259853 | 0.445839 | 0.016805 | 0.001416 | -0.10365 |
| das_8 | DAS | I plan my days activities in advance | 0.520493 | -0.12765 | 0.04413 | 0.179174 | -0.00298 |
| das_13 | DAS | I set goals for myself | 0.574001 | 0.081485 | -0.04016 | 0.074428 | -0.05929 |
| das_17 | DAS | When doing a demanding task, I have difficulty working out what I have to do | 0.095218 | 0.111098 | -0.07499 | -0.14177 | 0.549901 |
| ami_2 | AMI | I start conversations with random people | 0.11743 | -0.06268 | 0.36961 | -0.01009 | -0.09121 |
| ami_9 | AMI | When I decide to do something, I am able to make an effort easily | 0.54028 | -0.15503 | 0.225533 | -0.07572 | 0.235536 |
| ami_10 | AMI | I don't like to laze around | 0.592719 | -0.12753 | 0.031221 | -0.06747 | 0.034135 |
| ami_11 | AMI | I get things done when they need to be done, without requiring reminders from others | 0.640027 | -0.17306 | 0.005171 | 0.016291 | 0.315166 |
| ami_14 | AMI | I start conversations without being prompted | 0.082206 | 0.015403 | 0.451437 | 0.058664 | 0.059648 |
| ami_15 | AMI | When I have something I need to do, I do it straightaway so it is out of the way | 0.783389 | -0.26891 | -0.08525 | -0.02724 | 0.18936 |
| aes_2 | AES | I get things done during the day. | 0.490825 | 0.216379 | -0.07946 | 0.016603 | 0.269721 |
| aes_3 | AES | Getting things started on my own is important to me. | 0.318319 | 0.342016 | -0.1117 | -0.0542 | 0.069631 |
| aes_6 | AES | I put little effort into anything. | 0.127752 | 0.126977 | -0.09691 | 0.141168 | 0.360575 |
| aes_10 | AES | Someone has to tell me what to do each day. | 0.080953 | 0.152891 | -0.1576 | 0.089117 | 0.327242 |
| aes_16 | AES | Getting things done during the day is important to me. | 0.498339 | 0.241264 | -0.09394 | 0.147427 | 0.04793 |
| aes_17 | AES | I have initiative. | 0.461976 | 0.424351 | -0.01502 | -0.03576 | 0.045615 |
| das_1 | DAS | I need a bit of encouragement to get things started | 0.36484 | -0.01967 | -0.04304 | -0.13826 | 0.458408 |
| das_2 | DAS | I contact my friends | 0.041979 | -0.12095 | 0.803103 | -0.02828 | 0.048061 |
| das_6 | DAS | I find myself staring in to space | 0.041995 | -0.01034 | 0.028721 | -0.06821 | 0.537035 |
| das_16 | DAS | I act on things I have thought about during the day | 0.510231 | 0.083331 | -0.02517 | 0.033801 | -0.15793 |
| das_18 | DAS | I keep myself busy | 0.499714 | 0.098877 | -0.08052 | 0.096978 | 0.036078 |
| das_22 | DAS | I am spontaneous | 0.062492 | 0.277375 | 0.23262 | -0.07238 | -0.27613 |
| ami_12 | AMI | When I decide to do something, I am motivated to see it through to the end | 0.709954 | -0.12275 | -0.018 | -0.00461 | 0.219735 |
| aes_8 | AES | Seeing a job through to the end is important to me. | 0.421366 | 0.309716 | -0.20506 | 0.108946 | 0.163558 |
| aes_9 | AES | I spend time doing things that interest me. | -0.0427 | 0.6475 | -0.00674 | -0.13994 | 0.083566 |
| das_10 | DAS | I am able to focus on a task until it is finished | 0.54307 | -0.01398 | -0.11868 | 0.033454 | 0.355552 |
| das_19 | DAS | I get easily confused when doing several things at once | 0.001116 | 0.107716 | 0.165978 | -0.133 | 0.507139 |
| das_21 | DAS | I find it difficult to keep my mind on things | 0.244821 | -0.009 | 0.063477 | -0.07837 | 0.656031 |
| das_23 | DAS | I am easily distracted | 0.412838 | -0.19678 | 0.024424 | -0.10081 | 0.59796 |
| ami_1 | AMI | I feel sad or upset when I hear bad news | 0.014451 | -0.00635 | -0.06332 | 0.557456 | -0.15994 |
| ami_7 | AMI | Based on the last two weeks, I would say I care deeply about how my loved ones think of me | 0.047364 | -0.04134 | 0.11673 | 0.438543 | 0.025813 |
| ami_16 | AMI | I feel bad when I hear an acquaintance has an accident or illness | 0.013028 | 0.047459 | -0.02663 | 0.561374 | 0.030043 |
| aes_14 | AES | When something good happens, I get excited. | -0.01934 | 0.412249 | 0.226915 | 0.163938 | -0.10904 |
| das_3 | DAS | I express my emotions | 0.164648 | 0.009504 | 0.295819 | 0.243126 | 0.004815 |
| das_9 | DAS | When I receive bad news I feel bad about it | 0.040984 | 0.012172 | -0.08976 | 0.598041 | -0.13931 |
| das_12 | DAS | I struggle to empathise with other people | -0.21736 | 0.161304 | 0.102593 | 0.388551 | 0.396928 |
| das_20 | DAS | I become emotional easily when watching something happy or sad on TV | 0.091092 | -0.07168 | -0.05434 | 0.456474 | -0.23425 |
| ami_6 | AMI | After making a decision, I will wonder if I have made the wrong choice | -0.13242 | -0.05503 | -0.0564 | 0.412522 | -0.1539 |
| ami_13 | AMI | I feel awful if I say something insensitive | 0.015687 | -0.13493 | 0.013921 | 0.719404 | -0.02322 |
| ami_18 | AMI | If I realise I have been unpleasant to someone, I will feel terribly guilty afterwards | -0.05376 | 0.052597 | -0.09726 | 0.730253 | -0.02929 |
| aes_11 | AES | I am less concerned about my problems than I should be. | 0.081155 | -0.13839 | 0.064097 | 0.203454 | 0.340813 |
| das_7 | DAS | Before I do something I think about how others would feel about it | 0.126108 | -0.07166 | 0.12672 | 0.395721 | -0.05252 |
| das_15 | DAS | I am unconcerned about how others feel about my behaviour | -0.17375 | -0.053 | 0.025927 | 0.43341 | 0.24417 |
| aes_15 | AES | I have an accurate understanding of my problems. | 0.092109 | 0.314548 | 0.028452 | 0.084558 | 0.087225 |
| aes_18 | AES | I have motivation. | 0.544078 | 0.32753 | 0.020882 | -0.01976 | 0.097048 |
| das_11 | DAS | I lack motivation | 0.456626 | 0.079402 | 0.065205 | -0.06516 | 0.441757 |

**Supplementary Figure 1:**
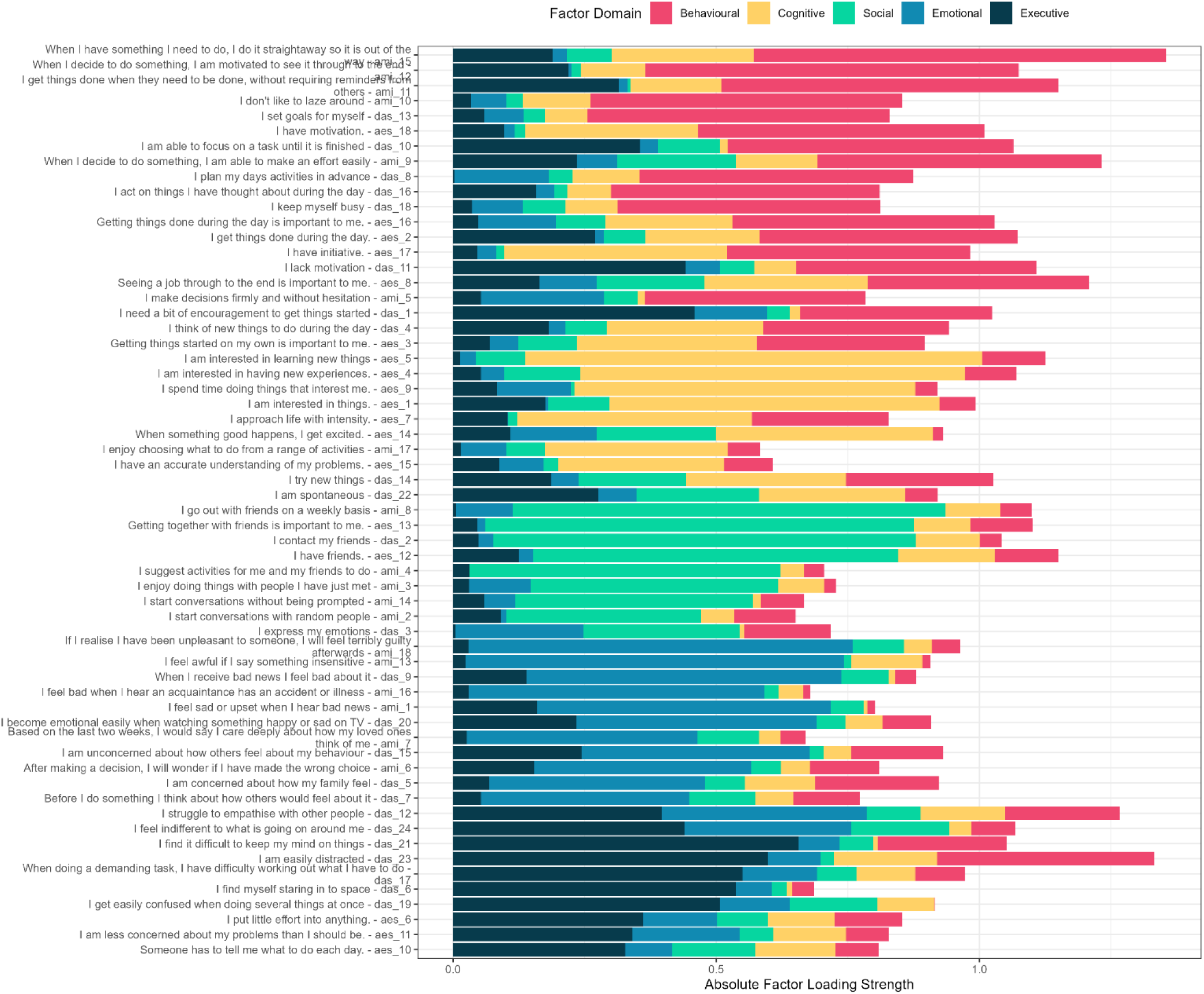
Stacked loading plot.

**Supplementary Table 2:** Module characteristics for the HC Cohort from 16 to 90. This table details the properties of network modules identified across different age windows, where each row represents a single module. The columns describe the following: Age is the centre of the 10-year sliding age window (in years); ModuleID is a unique identifier for each module within that age window; CentralNode is the most influential node with the highest connection strength within its module; CentralNodeDomain is the pre-defined functional domain of that central node; ModuleSize indicates the total number of nodes in the module; DominantDomain is the most frequent functional domain among all nodes within the module; Purity measures module homogeneity as the proportion of nodes belonging to the dominant domain; and Entropy measures module diversity, with higher values indicating a greater mix of functional domains.

| Age | ModuleID | CentralNode | CentralNodeDomain | ModuleSize | DominantDomain | Purity | Entropy |
| --- | --- | --- | --- | --- | --- | --- | --- |
| 16 | 1 | ami_15 | Behavioural | 6 | Behavioural | 1 | 0 |
| 16 | 2 | ami_14 | Social | 6 | Social | 1 | 0 |
| 16 | 3 | ami_18 | Emotional | 6 | Emotional | 1 | 0 |
| 17 | 1 | ami_15 | Behavioural | 6 | Behavioural | 1 | 0 |
| 17 | 2 | ami_14 | Social | 6 | Social | 1 | 0 |
| 17 | 3 | ami_18 | Emotional | 6 | Emotional | 1 | 0 |
| 18 | 1 | ami_15 | Behavioural | 6 | Behavioural | 1 | 0 |
| 18 | 2 | ami_14 | Social | 6 | Social | 1 | 0 |
| 18 | 3 | ami_18 | Emotional | 6 | Emotional | 1 | 0 |
| 19 | 1 | ami_15 | Behavioural | 6 | Behavioural | 1 | 0 |
| 19 | 2 | ami_14 | Social | 6 | Social | 1 | 0 |
| 19 | 3 | ami_18 | Emotional | 6 | Emotional | 1 | 0 |
| 20 | 1 | ami_15 | Behavioural | 6 | Behavioural | 1 | 0 |
| 20 | 2 | ami_14 | Social | 6 | Social | 1 | 0 |
| 20 | 3 | ami_18 | Emotional | 6 | Emotional | 1 | 0 |
| 21 | 1 | ami_15 | Behavioural | 6 | Behavioural | 1 | 0 |
| 21 | 2 | ami_14 | Social | 6 | Social | 1 | 0 |
| 21 | 3 | ami_18 | Emotional | 6 | Emotional | 1 | 0 |
| 22 | 1 | ami_11 | Behavioural | 6 | Behavioural | 1 | 0 |
| 22 | 2 | ami_3 | Social | 6 | Social | 1 | 0 |
| 22 | 3 | ami_18 | Emotional | 6 | Emotional | 1 | 0 |
| 23 | 1 | ami_15 | Behavioural | 7 | Behavioural | 0.857143 | 0.591673 |
| 23 | 2 | ami_3 | Social | 6 | Social | 1 | 0 |
| 23 | 3 | ami_18 | Emotional | 5 | Emotional | 1 | 0 |
| 24 | 1 | ami_15 | Behavioural | 7 | Behavioural | 0.857143 | 0.591673 |
| 24 | 2 | ami_3 | Social | 6 | Social | 1 | 0 |
| 24 | 3 | ami_18 | Emotional | 5 | Emotional | 1 | 0 |
| 25 | 1 | ami_11 | Behavioural | 7 | Behavioural | 0.857143 | 0.591673 |
| 25 | 2 | ami_3 | Social | 6 | Social | 1 | 0 |
| 25 | 3 | ami_18 | Emotional | 5 | Emotional | 1 | 0 |
| 26 | 1 | ami_11 | Behavioural | 7 | Behavioural | 0.857143 | 0.591673 |
| 26 | 2 | ami_3 | Social | 6 | Social | 1 | 0 |
| 26 | 3 | ami_13 | Emotional | 5 | Emotional | 1 | 0 |
| 27 | 1 | ami_11 | Behavioural | 7 | Behavioural | 0.857143 | 0.591673 |
| 27 | 2 | ami_3 | Social | 6 | Social | 1 | 0 |
| 27 | 3 | ami_13 | Emotional | 5 | Emotional | 1 | 0 |
| 28 | 1 | ami_15 | Behavioural | 7 | Behavioural | 0.857143 | 0.591673 |
| 28 | 2 | ami_3 | Social | 6 | Social | 1 | 0 |
| 28 | 3 | ami_18 | Emotional | 5 | Emotional | 1 | 0 |
| 29 | 1 | ami_15 | Behavioural | 7 | Behavioural | 0.857143 | 0.591673 |
| 29 | 2 | ami_3 | Social | 6 | Social | 1 | 0 |
| 29 | 3 | ami_18 | Emotional | 5 | Emotional | 1 | 0 |
| 30 | 1 | ami_3 | Social | 6 | Social | 1 | 0 |
| 30 | 2 | ami_18 | Emotional | 5 | Emotional | 1 | 0 |
| 30 | 3 | ami_15 | Behavioural | 7 | Behavioural | 0.857143 | 0.591673 |
| 31 | 1 | ami_15 | Behavioural | 7 | Behavioural | 0.857143 | 0.591673 |
| 31 | 2 | ami_3 | Social | 6 | Social | 1 | 0 |
| 31 | 3 | ami_13 | Emotional | 5 | Emotional | 1 | 0 |
| 32 | 1 | ami_3 | Social | 6 | Social | 1 | 0 |
| 32 | 2 | ami_13 | Emotional | 5 | Emotional | 1 | 0 |
| 32 | 3 | ami_15 | Behavioural | 7 | Behavioural | 0.857143 | 0.591673 |
| 33 | 1 | ami_15 | Behavioural | 7 | Behavioural | 0.857143 | 0.591673 |
| 33 | 2 | ami_3 | Social | 6 | Social | 1 | 0 |
| 33 | 3 | ami_18 | Emotional | 5 | Emotional | 1 | 0 |
| 34 | 1 | ami_15 | Behavioural | 7 | Behavioural | 0.857143 | 0.591673 |
| 34 | 2 | ami_3 | Social | 6 | Social | 1 | 0 |
| 34 | 3 | ami_18 | Emotional | 5 | Emotional | 1 | 0 |
| 35 | 1 | ami_15 | Behavioural | 7 | Behavioural | 0.857143 | 0.591673 |
| 35 | 2 | ami_3 | Social | 6 | Social | 1 | 0 |
| 35 | 3 | ami_18 | Emotional | 5 | Emotional | 1 | 0 |
| 36 | 1 | ami_15 | Behavioural | 7 | Behavioural | 0.857143 | 0.591673 |
| 36 | 2 | ami_3 | Social | 6 | Social | 1 | 0 |
| 36 | 3 | ami_13 | Emotional | 5 | Emotional | 1 | 0 |
| 37 | 1 | ami_15 | Behavioural | 7 | Behavioural | 0.857143 | 0.591673 |
| 37 | 2 | ami_3 | Social | 6 | Social | 1 | 0 |
| 37 | 3 | ami_13 | Emotional | 5 | Emotional | 1 | 0 |
| 38 | 1 | ami_11 | Behavioural | 7 | Behavioural | 0.857143 | 0.591673 |
| 38 | 2 | ami_3 | Social | 6 | Social | 1 | 0 |
| 38 | 3 | ami_13 | Emotional | 5 | Emotional | 1 | 0 |
| 39 | 1 | ami_11 | Behavioural | 7 | Behavioural | 0.857143 | 0.591673 |
| 39 | 2 | ami_3 | Social | 6 | Social | 1 | 0 |
| 39 | 3 | ami_13 | Emotional | 5 | Emotional | 1 | 0 |
| 40 | 1 | ami_11 | Behavioural | 7 | Behavioural | 0.857143 | 0.591673 |
| 40 | 2 | ami_4 | Social | 6 | Social | 1 | 0 |
| 40 | 3 | ami_13 | Emotional | 5 | Emotional | 1 | 0 |
| 41 | 1 | ami_11 | Behavioural | 7 | Behavioural | 0.857143 | 0.591673 |
| 41 | 2 | ami_4 | Social | 6 | Social | 1 | 0 |
| 41 | 3 | ami_13 | Emotional | 5 | Emotional | 1 | 0 |
| 42 | 1 | ami_11 | Behavioural | 7 | Behavioural | 0.857143 | 0.591673 |
| 42 | 2 | ami_4 | Social | 6 | Social | 1 | 0 |
| 42 | 3 | ami_13 | Emotional | 5 | Emotional | 1 | 0 |
| 43 | 1 | ami_11 | Behavioural | 7 | Behavioural | 0.857143 | 0.591673 |
| 43 | 2 | ami_4 | Social | 6 | Social | 1 | 0 |
| 43 | 3 | ami_13 | Emotional | 5 | Emotional | 1 | 0 |
| 44 | 1 | ami_11 | Behavioural | 7 | Behavioural | 0.857143 | 0.591673 |
| 44 | 2 | ami_4 | Social | 6 | Social | 1 | 0 |
| 44 | 3 | ami_13 | Emotional | 5 | Emotional | 1 | 0 |
| 45 | 1 | ami_11 | Behavioural | 7 | Behavioural | 0.857143 | 0.591673 |
| 45 | 2 | ami_4 | Social | 6 | Social | 1 | 0 |
| 45 | 3 | ami_18 | Emotional | 5 | Emotional | 1 | 0 |
| 46 | 1 | ami_11 | Behavioural | 7 | Behavioural | 0.857143 | 0.591673 |
| 46 | 2 | ami_3 | Social | 6 | Social | 1 | 0 |
| 46 | 3 | ami_18 | Emotional | 5 | Emotional | 1 | 0 |
| 47 | 1 | ami_11 | Behavioural | 7 | Behavioural | 0.857143 | 0.591673 |
| 47 | 2 | ami_3 | Social | 6 | Social | 1 | 0 |
| 47 | 3 | ami_18 | Emotional | 5 | Emotional | 1 | 0 |
| 48 | 1 | ami_12 | Behavioural | 7 | Behavioural | 0.857143 | 0.591673 |
| 48 | 2 | ami_3 | Social | 6 | Social | 1 | 0 |
| 48 | 3 | ami_18 | Emotional | 5 | Emotional | 1 | 0 |
| 49 | 1 | ami_12 | Behavioural | 7 | Behavioural | 0.857143 | 0.591673 |
| 49 | 2 | ami_2 | Social | 3 | Social | 1 | 0 |
| 49 | 3 | ami_4 | Social | 3 | Social | 1 | 0 |
| 49 | 4 | ami_18 | Emotional | 5 | Emotional | 1 | 0 |
| 50 | 1 | ami_12 | Behavioural | 7 | Behavioural | 0.857143 | 0.591673 |
| 50 | 2 | ami_2 | Social | 3 | Social | 1 | 0 |
| 50 | 3 | ami_4 | Social | 3 | Social | 1 | 0 |
| 50 | 4 | ami_18 | Emotional | 5 | Emotional | 1 | 0 |
| 51 | 1 | ami_12 | Behavioural | 7 | Behavioural | 0.857143 | 0.591673 |
| 51 | 2 | ami_2 | Social | 3 | Social | 1 | 0 |
| 51 | 3 | ami_4 | Social | 3 | Social | 1 | 0 |
| 51 | 4 | ami_18 | Emotional | 5 | Emotional | 1 | 0 |
| 52 | 1 | ami_3 | Social | 6 | Social | 1 | 0 |
| 52 | 2 | ami_13 | Emotional | 5 | Emotional | 1 | 0 |
| 52 | 3 | ami_12 | Behavioural | 7 | Behavioural | 0.857143 | 0.591673 |
| 53 | 1 | ami_12 | Behavioural | 7 | Behavioural | 0.857143 | 0.591673 |
| 53 | 2 | ami_3 | Social | 6 | Social | 1 | 0 |
| 53 | 3 | ami_13 | Emotional | 5 | Emotional | 1 | 0 |
| 54 | 1 | ami_3 | Social | 6 | Social | 1 | 0 |
| 54 | 2 | ami_13 | Emotional | 5 | Emotional | 1 | 0 |
| 54 | 3 | ami_12 | Behavioural | 7 | Behavioural | 0.857143 | 0.591673 |
| 55 | 1 | ami_12 | Behavioural | 7 | Behavioural | 0.857143 | 0.591673 |
| 55 | 2 | ami_3 | Social | 6 | Social | 1 | 0 |
| 55 | 3 | ami_13 | Emotional | 5 | Emotional | 1 | 0 |
| 56 | 1 | ami_12 | Behavioural | 7 | Behavioural | 0.857143 | 0.591673 |
| 56 | 2 | ami_4 | Social | 6 | Social | 1 | 0 |
| 56 | 3 | ami_13 | Emotional | 5 | Emotional | 1 | 0 |
| 57 | 1 | ami_12 | Behavioural | 7 | Behavioural | 0.857143 | 0.591673 |
| 57 | 2 | ami_4 | Social | 6 | Social | 1 | 0 |
| 57 | 3 | ami_13 | Emotional | 5 | Emotional | 1 | 0 |
| 58 | 1 | ami_12 | Behavioural | 6 | Behavioural | 1 | 0 |
| 58 | 2 | ami_4 | Social | 6 | Social | 1 | 0 |
| 58 | 3 | ami_13 | Emotional | 6 | Emotional | 1 | 0 |
| 59 | 1 | ami_12 | Behavioural | 6 | Behavioural | 1 | 0 |
| 59 | 2 | ami_4 | Social | 6 | Social | 1 | 0 |
| 59 | 3 | ami_13 | Emotional | 6 | Emotional | 1 | 0 |
| 60 | 1 | ami_12 | Behavioural | 6 | Behavioural | 1 | 0 |
| 60 | 2 | ami_4 | Social | 6 | Social | 1 | 0 |
| 60 | 3 | ami_13 | Emotional | 6 | Emotional | 1 | 0 |
| 61 | 1 | ami_12 | Behavioural | 7 | Behavioural | 0.857143 | 0.591673 |
| 61 | 2 | ami_4 | Social | 5 | Social | 1 | 0 |
| 61 | 3 | ami_13 | Emotional | 6 | Emotional | 1 | 0 |
| 62 | 1 | ami_12 | Behavioural | 8 | Behavioural | 0.75 | 0.811278 |
| 62 | 2 | ami_3 | Social | 2 | Social | 1 | 0 |
| 62 | 3 | ami_4 | Social | 2 | Social | 1 | 0 |
| 62 | 4 | ami_13 | Emotional | 6 | Emotional | 1 | 0 |
| 63 | 1 | ami_12 | Behavioural | 8 | Behavioural | 0.75 | 0.811278 |
| 63 | 2 | ami_3 | Social | 2 | Social | 1 | 0 |
| 63 | 3 | ami_4 | Social | 2 | Social | 1 | 0 |
| 63 | 4 | ami_13 | Emotional | 6 | Emotional | 1 | 0 |
| 64 | 1 | ami_12 | Behavioural | 8 | Behavioural | 0.75 | 0.811278 |
| 64 | 2 | ami_3 | Social | 2 | Social | 1 | 0 |
| 64 | 3 | ami_4 | Social | 2 | Social | 1 | 0 |
| 64 | 4 | ami_13 | Emotional | 6 | Emotional | 1 | 0 |
| 65 | 1 | ami_12 | Behavioural | 8 | Behavioural | 0.75 | 0.811278 |
| 65 | 2 | ami_3 | Social | 2 | Social | 1 | 0 |
| 65 | 3 | ami_4 | Social | 2 | Social | 1 | 0 |
| 65 | 4 | ami_18 | Emotional | 6 | Emotional | 1 | 0 |
| 66 | 1 | ami_12 | Behavioural | 8 | Behavioural | 0.75 | 0.811278 |
| 66 | 2 | ami_3 | Social | 2 | Social | 1 | 0 |
| 66 | 3 | ami_4 | Social | 2 | Social | 1 | 0 |
| 66 | 4 | ami_18 | Emotional | 6 | Emotional | 1 | 0 |
| 67 | 1 | ami_12 | Behavioural | 8 | Behavioural | 0.75 | 0.811278 |
| 67 | 2 | ami_3 | Social | 2 | Social | 1 | 0 |
| 67 | 3 | ami_4 | Social | 2 | Social | 1 | 0 |
| 67 | 4 | ami_18 | Emotional | 6 | Emotional | 1 | 0 |
| 68 | 1 | ami_12 | Behavioural | 8 | Behavioural | 0.75 | 0.811278 |
| 68 | 2 | ami_3 | Social | 2 | Social | 1 | 0 |
| 68 | 3 | ami_4 | Social | 2 | Social | 1 | 0 |
| 68 | 4 | ami_18 | Emotional | 6 | Emotional | 1 | 0 |
| 69 | 1 | ami_12 | Behavioural | 8 | Behavioural | 0.75 | 0.811278 |
| 69 | 2 | ami_3 | Social | 2 | Social | 1 | 0 |
| 69 | 3 | ami_4 | Social | 2 | Social | 1 | 0 |
| 69 | 4 | ami_18 | Emotional | 6 | Emotional | 1 | 0 |
| 70 | 1 | ami_12 | Behavioural | 8 | Behavioural | 0.75 | 0.811278 |
| 70 | 2 | ami_3 | Social | 2 | Social | 1 | 0 |
| 70 | 3 | ami_4 | Social | 2 | Social | 1 | 0 |
| 70 | 4 | ami_18 | Emotional | 6 | Emotional | 1 | 0 |
| 71 | 1 | ami_12 | Behavioural | 7 | Behavioural | 0.857143 | 0.591673 |
| 71 | 2 | ami_3 | Social | 2 | Social | 1 | 0 |
| 71 | 3 | ami_4 | Social | 3 | Social | 1 | 0 |
| 71 | 4 | ami_18 | Emotional | 6 | Emotional | 1 | 0 |
| 72 | 1 | ami_12 | Behavioural | 8 | Behavioural | 0.75 | 0.811278 |
| 72 | 2 | ami_3 | Social | 2 | Social | 1 | 0 |
| 72 | 3 | ami_4 | Social | 2 | Social | 1 | 0 |
| 72 | 4 | ami_18 | Emotional | 6 | Emotional | 1 | 0 |
| 73 | 1 | ami_12 | Behavioural | 8 | Behavioural | 0.75 | 0.811278 |
| 73 | 2 | ami_3 | Social | 2 | Social | 1 | 0 |
| 73 | 3 | ami_4 | Social | 2 | Social | 1 | 0 |
| 73 | 4 | ami_18 | Emotional | 6 | Emotional | 1 | 0 |
| 74 | 1 | ami_12 | Behavioural | 8 | Behavioural | 0.75 | 0.811278 |
| 74 | 2 | ami_4 | Social | 4 | Social | 1 | 0 |
| 74 | 3 | ami_18 | Emotional | 6 | Emotional | 1 | 0 |
| 75 | 1 | ami_11 | Behavioural | 8 | Behavioural | 0.75 | 0.811278 |
| 75 | 2 | ami_4 | Social | 6 | Social | 0.666667 | 0.918296 |
| 75 | 3 | ami_18 | Emotional | 4 | Emotional | 1 | 0 |
| 76 | 1 | ami_12 | Behavioural | 8 | Behavioural | 0.75 | 0.811278 |
| 76 | 2 | ami_4 | Social | 6 | Social | 0.666667 | 0.918296 |
| 76 | 3 | ami_18 | Emotional | 4 | Emotional | 1 | 0 |
| 77 | 1 | ami_12 | Behavioural | 9 | Behavioural | 0.666667 | 1.224394 |
| 77 | 2 | ami_4 | Social | 6 | Social | 0.666667 | 0.918296 |
| 77 | 3 | ami_13 | Emotional | 3 | Emotional | 1 | 0 |
| 78 | 1 | ami_12 | Behavioural | 9 | Behavioural | 0.666667 | 1.224394 |
| 78 | 2 | ami_4 | Social | 6 | Social | 0.666667 | 0.918296 |
| 78 | 3 | ami_13 | Emotional | 3 | Emotional | 1 | 0 |
| 79 | 1 | ami_12 | Behavioural | 8 | Behavioural | 0.75 | 0.811278 |
| 79 | 2 | ami_4 | Social | 6 | Social | 0.666667 | 0.918296 |
| 79 | 3 | ami_16 | Emotional | 4 | Emotional | 1 | 0 |
| 80 | 1 | ami_12 | Behavioural | 10 | Behavioural | 0.6 | 1.370951 |
| 80 | 2 | ami_4 | Social | 5 | Social | 0.8 | 0.721928 |
| 80 | 3 | ami_13 | Emotional | 3 | Emotional | 1 | 0 |
| 81 | 1 | ami_11 | Behavioural | 9 | Behavioural | 0.666667 | 1.224394 |
| 81 | 2 | ami_16 | Emotional | 4 | Emotional | 1 | 0 |
| 81 | 3 | ami_4 | Social | 5 | Social | 0.8 | 0.721928 |
| 82 | 1 | ami_12 | Behavioural | 10 | Behavioural | 0.6 | 1.370951 |
| 82 | 2 | ami_2 | Social | 4 | Social | 1 | 0 |
| 82 | 3 | ami_18 | Emotional | 4 | Emotional | 1 | 0 |
| 83 | 1 | ami_9 | Behavioural | 6 | Behavioural | 0.666667 | 1.251629 |
| 83 | 2 | ami_2 | Social | 4 | Social | 1 | 0 |
| 83 | 3 | ami_12 | Behavioural | 8 | Emotional | 0.625 | 1.298795 |
| 84 | 1 | ami_2 | Social | 7 | Social | 0.714286 | 1.148835 |
| 84 | 2 | ami_12 | Behavioural | 11 | Behavioural | 0.454545 | 1.348588 |
| 85 | 1 | ami_9 | Behavioural | 3 | Behavioural | 0.666667 | 0.918296 |
| 85 | 2 | ami_2 | Social | 6 | Social | 0.833333 | 0.650022 |
| 85 | 3 | ami_12 | Behavioural | 9 | Emotional | 0.555556 | 1.351644 |

**Supplementary Table 3:** Module characteristics for the Depression cohort.

| Age | ModuleID | CentralNode | CentralNodeDomain | ModuleSize | DominantDomain | Purity | Entropy |
| --- | --- | --- | --- | --- | --- | --- | --- |
| 16 | 1 | ami_12 | Behavioural | 6 | Behavioural | 1 | 0 |
| 16 | 2 | ami_2 | Social | 5 | Social | 1 | 0 |
| 16 | 3 | ami_18 | Emotional | 7 | Emotional | 0.857143 | 0.591673 |
| 17 | 1 | ami_12 | Behavioural | 6 | Behavioural | 1 | 0 |
| 17 | 2 | ami_2 | Social | 5 | Social | 1 | 0 |
| 17 | 3 | ami_18 | Emotional | 7 | Emotional | 0.857143 | 0.591673 |
| 18 | 1 | ami_12 | Behavioural | 6 | Behavioural | 1 | 0 |
| 18 | 2 | ami_2 | Social | 5 | Social | 1 | 0 |
| 18 | 3 | ami_18 | Emotional | 7 | Emotional | 0.857143 | 0.591673 |
| 19 | 1 | ami_12 | Behavioural | 6 | Behavioural | 1 | 0 |
| 19 | 2 | ami_3 | Social | 6 | Social | 1 | 0 |
| 19 | 3 | ami_13 | Emotional | 6 | Emotional | 1 | 0 |
| 20 | 1 | ami_12 | Behavioural | 6 | Behavioural | 1 | 0 |
| 20 | 2 | ami_2 | Social | 3 | Social | 1 | 0 |
| 20 | 3 | ami_4 | Social | 3 | Social | 1 | 0 |
| 20 | 4 | ami_13 | Emotional | 6 | Emotional | 1 | 0 |
| 21 | 1 | ami_12 | Behavioural | 6 | Behavioural | 1 | 0 |
| 21 | 2 | ami_2 | Social | 3 | Social | 1 | 0 |
| 21 | 3 | ami_4 | Social | 3 | Social | 1 | 0 |
| 21 | 4 | ami_18 | Emotional | 6 | Emotional | 1 | 0 |
| 22 | 1 | ami_12 | Behavioural | 5 | Behavioural | 1 | 0 |
| 22 | 2 | ami_3 | Social | 4 | Social | 0.75 | 0.811278 |
| 22 | 3 | ami_4 | Social | 3 | Social | 1 | 0 |
| 22 | 4 | ami_18 | Emotional | 6 | Emotional | 1 | 0 |
| 23 | 1 | ami_12 | Behavioural | 6 | Behavioural | 0.833333 | 0.650022 |
| 23 | 2 | ami_5 | Behavioural | 2 | Behavioural | 0.5 | 1 |
| 23 | 3 | ami_3 | Social | 2 | Social | 1 | 0 |
| 23 | 4 | ami_4 | Social | 3 | Social | 1 | 0 |
| 23 | 5 | ami_13 | Emotional | 5 | Emotional | 1 | 0 |
| 24 | 1 | ami_12 | Behavioural | 5 | Behavioural | 1 | 0 |
| 24 | 2 | ami_5 | Behavioural | 3 | Behavioural | 0.333333 | 1.584963 |
| 24 | 3 | ami_3 | Social | 2 | Social | 1 | 0 |
| 24 | 4 | ami_4 | Social | 3 | Social | 1 | 0 |
| 24 | 5 | ami_13 | Emotional | 5 | Emotional | 1 | 0 |
| 25 | 1 | ami_12 | Behavioural | 5 | Behavioural | 1 | 0 |
| 25 | 2 | ami_14 | Social | 8 | Social | 0.75 | 1.061278 |
| 25 | 3 | ami_13 | Emotional | 5 | Emotional | 1 | 0 |
| 26 | 1 | ami_12 | Behavioural | 5 | Behavioural | 1 | 0 |
| 26 | 2 | ami_14 | Social | 8 | Social | 0.75 | 1.061278 |
| 26 | 3 | ami_13 | Emotional | 5 | Emotional | 1 | 0 |
| 27 | 1 | ami_12 | Behavioural | 5 | Behavioural | 1 | 0 |
| 27 | 2 | ami_4 | Social | 8 | Social | 0.75 | 1.061278 |
| 27 | 3 | ami_13 | Emotional | 5 | Emotional | 1 | 0 |
| 28 | 1 | ami_4 | Social | 5 | Social | 1 | 0 |
| 28 | 2 | ami_13 | Emotional | 5 | Emotional | 1 | 0 |
| 28 | 3 | ami_12 | Behavioural | 8 | Behavioural | 0.75 | 1.061278 |
| 29 | 1 | ami_12 | Behavioural | 8 | Behavioural | 0.75 | 1.061278 |
| 29 | 2 | ami_4 | Social | 5 | Social | 1 | 0 |
| 29 | 3 | ami_13 | Emotional | 5 | Emotional | 1 | 0 |
| 30 | 1 | ami_12 | Behavioural | 8 | Behavioural | 0.75 | 1.061278 |
| 30 | 2 | ami_4 | Social | 5 | Social | 1 | 0 |
| 30 | 3 | ami_13 | Emotional | 5 | Emotional | 1 | 0 |
| 31 | 1 | ami_12 | Behavioural | 8 | Behavioural | 0.75 | 1.061278 |
| 31 | 2 | ami_4 | Social | 5 | Social | 1 | 0 |
| 31 | 3 | ami_13 | Emotional | 5 | Emotional | 1 | 0 |
| 32 | 1 | ami_12 | Behavioural | 8 | Behavioural | 0.75 | 1.061278 |
| 32 | 2 | ami_4 | Social | 5 | Social | 1 | 0 |
| 32 | 3 | ami_13 | Emotional | 5 | Emotional | 1 | 0 |
| 33 | 1 | ami_12 | Behavioural | 8 | Behavioural | 0.75 | 1.061278 |
| 33 | 2 | ami_4 | Social | 5 | Social | 1 | 0 |
| 33 | 3 | ami_13 | Emotional | 5 | Emotional | 1 | 0 |
| 34 | 1 | ami_15 | Behavioural | 3 | Behavioural | 1 | 0 |
| 34 | 2 | ami_4 | Social | 5 | Social | 1 | 0 |
| 34 | 3 | ami_13 | Emotional | 5 | Emotional | 1 | 0 |
| 34 | 4 | ami_5 | Behavioural | 5 | Behavioural | 0.6 | 1.370951 |
| 35 | 1 | ami_15 | Behavioural | 3 | Behavioural | 1 | 0 |
| 35 | 2 | ami_5 | Behavioural | 5 | Behavioural | 0.6 | 1.370951 |
| 35 | 3 | ami_4 | Social | 5 | Social | 1 | 0 |
| 35 | 4 | ami_13 | Emotional | 5 | Emotional | 1 | 0 |
| 36 | 1 | ami_15 | Behavioural | 3 | Behavioural | 1 | 0 |
| 36 | 2 | ami_5 | Behavioural | 5 | Behavioural | 0.6 | 1.370951 |
| 36 | 3 | ami_4 | Social | 5 | Social | 1 | 0 |
| 36 | 4 | ami_18 | Emotional | 5 | Emotional | 1 | 0 |
| 37 | 1 | ami_15 | Behavioural | 3 | Behavioural | 1 | 0 |
| 37 | 2 | ami_5 | Behavioural | 5 | Behavioural | 0.6 | 1.370951 |
| 37 | 3 | ami_4 | Social | 5 | Social | 1 | 0 |
| 37 | 4 | ami_18 | Emotional | 5 | Emotional | 1 | 0 |
| 38 | 1 | ami_15 | Behavioural | 3 | Behavioural | 1 | 0 |
| 38 | 2 | ami_5 | Behavioural | 5 | Behavioural | 0.6 | 1.370951 |
| 38 | 3 | ami_4 | Social | 5 | Social | 1 | 0 |
| 38 | 4 | ami_18 | Emotional | 5 | Emotional | 1 | 0 |
| 39 | 1 | ami_15 | Behavioural | 3 | Behavioural | 1 | 0 |
| 39 | 2 | ami_5 | Behavioural | 5 | Behavioural | 0.6 | 1.370951 |
| 39 | 3 | ami_4 | Social | 5 | Social | 1 | 0 |
| 39 | 4 | ami_18 | Emotional | 5 | Emotional | 1 | 0 |
| 40 | 1 | ami_15 | Behavioural | 3 | Behavioural | 1 | 0 |
| 40 | 2 | ami_5 | Behavioural | 5 | Behavioural | 0.6 | 1.370951 |
| 40 | 3 | ami_2 | Social | 3 | Social | 1 | 0 |
| 40 | 4 | ami_4 | Social | 2 | Social | 1 | 0 |
| 40 | 5 | ami_18 | Emotional | 3 | Emotional | 1 | 0 |
| 40 | 6 | ami_16 | Emotional | 2 | Emotional | 1 | 0 |
| 41 | 1 | ami_15 | Behavioural | 3 | Behavioural | 1 | 0 |
| 41 | 2 | ami_9 | Behavioural | 5 | Behavioural | 0.6 | 1.370951 |
| 41 | 3 | ami_4 | Social | 5 | Social | 1 | 0 |
| 41 | 4 | ami_18 | Emotional | 5 | Emotional | 1 | 0 |
| 42 | 1 | ami_15 | Behavioural | 5 | Behavioural | 1 | 0 |
| 42 | 2 | ami_5 | Behavioural | 2 | Behavioural | 0.5 | 1 |
| 42 | 3 | ami_2 | Social | 3 | Social | 1 | 0 |
| 42 | 4 | ami_4 | Social | 3 | Social | 1 | 0 |
| 42 | 5 | ami_13 | Emotional | 5 | Emotional | 1 | 0 |
| 43 | 1 | ami_15 | Behavioural | 3 | Behavioural | 1 | 0 |
| 43 | 2 | ami_2 | Social | 3 | Social | 1 | 0 |
| 43 | 3 | ami_4 | Social | 3 | Social | 1 | 0 |
| 43 | 4 | ami_13 | Emotional | 5 | Emotional | 1 | 0 |
| 43 | 5 | ami_9 | Behavioural | 4 | Behavioural | 0.75 | 0.811278 |
| 44 | 1 | ami_15 | Behavioural | 5 | Behavioural | 1 | 0 |
| 44 | 2 | ami_5 | Behavioural | 2 | Behavioural | 0.5 | 1 |
| 44 | 3 | ami_2 | Social | 3 | Social | 1 | 0 |
| 44 | 4 | ami_4 | Social | 3 | Social | 1 | 0 |
| 44 | 5 | ami_18 | Emotional | 5 | Emotional | 1 | 0 |
| 45 | 1 | ami_15 | Behavioural | 5 | Behavioural | 1 | 0 |
| 45 | 2 | ami_2 | Social | 3 | Social | 1 | 0 |
| 45 | 3 | ami_4 | Social | 3 | Social | 1 | 0 |
| 45 | 4 | ami_18 | Emotional | 5 | Emotional | 1 | 0 |
| 45 | 5 | ami_5 | Behavioural | 2 | Behavioural | 0.5 | 1 |
| 46 | 1 | ami_15 | Behavioural | 6 | Behavioural | 0.666667 | 0.918296 |
| 46 | 2 | ami_2 | Social | 4 | Social | 1 | 0 |
| 46 | 3 | ami_16 | Emotional | 6 | Emotional | 0.833333 | 0.650022 |
| 46 | 4 | ami_5 | Behavioural | 2 | Behavioural | 0.5 | 1 |
| 47 | 1 | ami_15 | Behavioural | 5 | Behavioural | 1 | 0 |
| 47 | 2 | ami_2 | Social | 7 | Social | 0.857143 | 0.591673 |
| 47 | 3 | ami_13 | Emotional | 6 | Emotional | 1 | 0 |
| 48 | 1 | ami_15 | Behavioural | 6 | Behavioural | 0.833333 | 0.650022 |
| 48 | 2 | ami_2 | Social | 5 | Social | 0.8 | 0.721928 |
| 48 | 3 | ami_13 | Emotional | 7 | Emotional | 0.857143 | 0.591673 |
| 49 | 1 | ami_12 | Behavioural | 8 | Behavioural | 0.625 | 1.298795 |
| 49 | 2 | ami_2 | Social | 4 | Social | 1 | 0 |
| 49 | 3 | ami_18 | Emotional | 2 | Emotional | 1 | 0 |
| 49 | 4 | ami_16 | Emotional | 2 | Emotional | 1 | 0 |
| 49 | 5 | ami_5 | Behavioural | 2 | Behavioural | 0.5 | 1 |
| 50 | 1 | ami_12 | Behavioural | 6 | Behavioural | 0.833333 | 0.650022 |
| 50 | 2 | ami_3 | Social | 4 | Social | 1 | 0 |
| 50 | 3 | ami_13 | Emotional | 6 | Emotional | 0.833333 | 0.650022 |
| 50 | 4 | ami_5 | Behavioural | 2 | Behavioural | 0.5 | 1 |
| 51 | 1 | ami_12 | Behavioural | 6 | Behavioural | 0.833333 | 0.650022 |
| 51 | 2 | ami_5 | Behavioural | 2 | Behavioural | 0.5 | 1 |
| 51 | 3 | ami_2 | Social | 4 | Social | 1 | 0 |
| 51 | 4 | ami_13 | Emotional | 6 | Emotional | 0.833333 | 0.650022 |
| 52 | 1 | ami_12 | Behavioural | 5 | Behavioural | 1 | 0 |
| 52 | 2 | ami_2 | Social | 5 | Social | 0.6 | 1.370951 |
| 52 | 3 | ami_4 | Social | 3 | Social | 1 | 0 |
| 52 | 4 | ami_18 | Emotional | 5 | Emotional | 1 | 0 |
| 53 | 1 | ami_12 | Behavioural | 5 | Behavioural | 1 | 0 |
| 53 | 2 | ami_2 | Social | 5 | Social | 0.6 | 1.370951 |
| 53 | 3 | ami_4 | Social | 3 | Social | 1 | 0 |
| 53 | 4 | ami_18 | Emotional | 5 | Emotional | 1 | 0 |
| 54 | 1 | ami_12 | Behavioural | 5 | Behavioural | 1 | 0 |
| 54 | 2 | ami_2 | Social | 5 | Social | 0.6 | 1.370951 |
| 54 | 3 | ami_4 | Social | 3 | Social | 1 | 0 |
| 54 | 4 | ami_18 | Emotional | 5 | Emotional | 1 | 0 |
| 55 | 1 | ami_12 | Behavioural | 5 | Behavioural | 1 | 0 |
| 55 | 2 | ami_5 | Behavioural | 2 | Behavioural | 0.5 | 1 |
| 55 | 3 | ami_4 | Social | 6 | Social | 1 | 0 |
| 55 | 4 | ami_18 | Emotional | 5 | Emotional | 1 | 0 |
| 56 | 1 | ami_12 | Behavioural | 6 | Behavioural | 0.833333 | 0.650022 |
| 56 | 2 | ami_5 | Behavioural | 2 | Behavioural | 0.5 | 1 |
| 56 | 3 | ami_4 | Social | 5 | Social | 1 | 0 |
| 56 | 4 | ami_18 | Emotional | 5 | Emotional | 1 | 0 |
| 57 | 1 | ami_15 | Behavioural | 5 | Behavioural | 1 | 0 |
| 57 | 2 | ami_3 | Social | 2 | Social | 1 | 0 |
| 57 | 3 | ami_4 | Social | 3 | Social | 1 | 0 |
| 57 | 4 | ami_18 | Emotional | 5 | Emotional | 1 | 0 |
| 57 | 5 | ami_5 | Behavioural | 3 | Behavioural | 0.333333 | 1.584963 |
| 58 | 1 | ami_15 | Behavioural | 7 | Behavioural | 0.857143 | 0.591673 |
| 58 | 2 | ami_3 | Social | 2 | Social | 1 | 0 |
| 58 | 3 | ami_4 | Social | 3 | Social | 1 | 0 |
| 58 | 4 | ami_18 | Emotional | 6 | Emotional | 1 | 0 |
| 59 | 1 | ami_12 | Behavioural | 7 | Behavioural | 0.857143 | 0.591673 |
| 59 | 2 | ami_4 | Social | 5 | Social | 1 | 0 |
| 59 | 3 | ami_18 | Emotional | 6 | Emotional | 1 | 0 |
| 60 | 1 | ami_15 | Behavioural | 8 | Behavioural | 0.75 | 1.061278 |
| 60 | 2 | ami_4 | Social | 5 | Social | 1 | 0 |
| 60 | 3 | ami_18 | Emotional | 5 | Emotional | 1 | 0 |
| 61 | 1 | ami_11 | Behavioural | 8 | Behavioural | 0.75 | 1.061278 |
| 61 | 2 | ami_3 | Social | 2 | Social | 1 | 0 |
| 61 | 3 | ami_8 | Social | 3 | Social | 0.666667 | 0.918296 |
| 61 | 4 | ami_18 | Emotional | 5 | Emotional | 0.8 | 0.721928 |
| 62 | 1 | ami_11 | Behavioural | 8 | Behavioural | 0.75 | 1.061278 |
| 62 | 2 | ami_4 | Social | 5 | Social | 0.8 | 0.721928 |
| 62 | 3 | ami_18 | Emotional | 5 | Emotional | 0.8 | 0.721928 |
| 63 | 1 | ami_11 | Behavioural | 7 | Behavioural | 0.714286 | 1.148835 |
| 63 | 2 | ami_3 | Social | 2 | Social | 1 | 0 |
| 63 | 3 | ami_4 | Social | 4 | Social | 0.5 | 1.5 |
| 63 | 4 | ami_18 | Emotional | 5 | Emotional | 0.8 | 0.721928 |
| 64 | 1 | ami_10 | Behavioural | 5 | Behavioural | 0.8 | 0.721928 |
| 64 | 2 | ami_3 | Social | 2 | Social | 1 | 0 |
| 64 | 3 | ami_9 | Behavioural | 5 | Behavioural | 0.4 | 1.521928 |
| 64 | 4 | ami_16 | Emotional | 6 | Emotional | 0.666667 | 0.918296 |
| 65 | 1 | ami_11 | Behavioural | 7 | Behavioural | 0.857143 | 0.591673 |
| 65 | 2 | ami_3 | Social | 3 | Social | 0.666667 | 0.918296 |
| 65 | 3 | ami_8 | Social | 3 | Social | 1 | 0 |
| 65 | 4 | ami_13 | Emotional | 5 | Emotional | 0.8 | 0.721928 |

**Supplementary Table 4:** Module characteristics for the Neurological Conditions cohort.

| Age | ModuleID | CentralNode | CentralNodeDomain | ModuleSize | DominantDomain | Purity | Entropy |
| --- | --- | --- | --- | --- | --- | --- | --- |
| 43 | 1 | ami_11 | Behavioural | 6 | Behavioural | 0.83333333 | 0.65002242 |
| 43 | 2 | ami_2 | Social | 3 | Social | 1 | 0 |
| 43 | 3 | ami_18 | Emotional | 5 | Emotional | 0.8 | 0.72192809 |
| 43 | 4 | ami_12 | Behavioural | 4 | Emotional | 0.5 | 1.5 |
| 44 | 1 | ami_12 | Behavioural | 7 | Emotional | 0.57142857 | 1.37878349 |
| 44 | 2 | ami_15 | Behavioural | 7 | Behavioural | 0.71428571 | 1.14883485 |
| 44 | 3 | ami_2 | Social | 4 | Social | 0.75 | 0.81127812 |
| 45 | 1 | ami_15 | Behavioural | 7 | Behavioural | 0.71428571 | 1.14883485 |
| 45 | 2 | ami_2 | Social | 4 | Social | 0.75 | 0.81127812 |
| 45 | 3 | ami_12 | Behavioural | 7 | Emotional | 0.57142857 | 1.37878349 |
| 46 | 1 | ami_15 | Behavioural | 7 | Behavioural | 0.71428571 | 1.14883485 |
| 46 | 2 | ami_2 | Social | 4 | Social | 1 | 0 |
| 46 | 3 | ami_16 | Emotional | 7 | Emotional | 0.71428571 | 1.14883485 |
| 47 | 1 | ami_15 | Behavioural | 8 | Behavioural | 0.75 | 1.06127812 |
| 47 | 2 | ami_14 | Social | 5 | Social | 1 | 0 |
| 47 | 3 | ami_18 | Emotional | 5 | Emotional | 1 | 0 |
| 48 | 1 | ami_15 | Behavioural | 7 | Behavioural | 0.85714286 | 0.59167278 |
| 48 | 2 | ami_3 | Social | 6 | Social | 1 | 0 |
| 48 | 3 | ami_18 | Emotional | 5 | Emotional | 1 | 0 |
| 49 | 1 | ami_9 | Behavioural | 8 | Behavioural | 0.75 | 1.06127812 |
| 49 | 2 | ami_3 | Social | 5 | Social | 1 | 0 |
| 49 | 3 | ami_18 | Emotional | 5 | Emotional | 1 | 0 |
| 50 | 1 | ami_12 | Behavioural | 7 | Behavioural | 0.85714286 | 0.59167278 |
| 50 | 2 | ami_3 | Social | 6 | Social | 1 | 0 |
| 50 | 3 | ami_18 | Emotional | 5 | Emotional | 1 | 0 |
| 51 | 1 | ami_12 | Behavioural | 7 | Behavioural | 0.85714286 | 0.59167278 |
| 51 | 2 | ami_3 | Social | 6 | Social | 1 | 0 |
| 51 | 3 | ami_18 | Emotional | 5 | Emotional | 1 | 0 |
| 52 | 1 | ami_12 | Behavioural | 5 | Behavioural | 1 | 0 |
| 52 | 2 | ami_3 | Social | 4 | Social | 0.75 | 0.81127812 |
| 52 | 3 | ami_4 | Social | 3 | Social | 1 | 0 |
| 52 | 4 | ami_18 | Emotional | 6 | Emotional | 1 | 0 |
| 53 | 1 | ami_12 | Behavioural | 7 | Behavioural | 0.71428571 | 0.86312057 |
| 53 | 2 | ami_3 | Social | 5 | Social | 0.8 | 0.72192809 |
| 53 | 3 | ami_18 | Emotional | 6 | Emotional | 1 | 0 |
| 54 | 1 | ami_12 | Behavioural | 7 | Behavioural | 0.7142857<br>1 | 0.8631205<br>7 |
| 54 | 2 | ami_3 | Social | 5 | Social | 0.8 | 0.7219280<br>9 |
| 54 | 3 | ami_18 | Emotional | 6 | Emotional | 1 | 0 |
| 55 | 1 | ami_12 | Behavioural | 6 | Behavioural | 0.8333333<br>3 | 0.6500224<br>2 |
| 55 | 2 | ami_4 | Social | 6 | Social | 0.8333333<br>3 | 0.6500224<br>2 |
| 55 | 3 | ami_18 | Emotional | 6 | Emotional | 1 | 0 |
| 56 | 1 | ami_12 | Behavioural | 6 | Behavioural | 0.8333333<br>3 | 0.6500224<br>2 |
| 56 | 2 | ami_4 | Social | 6 | Social | 0.8333333<br>3 | 0.6500224<br>2 |
| 56 | 3 | ami_18 | Emotional | 6 | Emotional | 1 | 0 |
| 57 | 1 | ami_12 | Behavioural | 6 | Behavioural | 0.8333333<br>3 | 0.6500224<br>2 |
| 57 | 2 | ami_4 | Social | 6 | Social | 0.8333333<br>3 | 0.6500224<br>2 |
| 57 | 3 | ami_18 | Emotional | 6 | Emotional | 1 | 0 |
| 58 | 1 | ami_12 | Behavioural | 6 | Behavioural | 0.8333333<br>3 | 0.6500224<br>2 |
| 58 | 2 | ami_4 | Social | 6 | Social | 0.8333333<br>3 | 0.6500224<br>2 |
| 58 | 3 | ami_18 | Emotional | 6 | Emotional | 1 | 0 |
| 59 | 1 | ami_12 | Behavioural | 6 | Behavioural | 0.8333333<br>3 | 0.6500224<br>2 |
| 59 | 2 | ami_4 | Social | 6 | Social | 0.8333333<br>3 | 0.6500224<br>2 |
| 59 | 3 | ami_18 | Emotional | 6 | Emotional | 1 | 0 |
| 60 | 1 | ami_12 | Behavioural | 9 | Behavioural | 0.6666666<br>7 | 1.2243944<br>5 |
| 60 | 2 | ami_4 | Social | 4 | Social | 1 | 0 |
| 60 | 3 | ami_18 | Emotional | 5 | Emotional | 1 | 0 |
| 61 | 1 | ami_12 | Behavioural | 9 | Behavioural | 0.6666666<br>7 | 1.2243944<br>5 |
| 61 | 2 | ami_4 | Social | 4 | Social | 1 | 0 |
| 61 | 3 | ami_18 | Emotional | 5 | Emotional | 1 | 0 |
| 62 | 1 | ami_12 | Behavioural | 9 | Behavioural | 0.6666666<br>7 | 1.2243944<br>5 |
| 62 | 2 | ami_4 | Social | 4 | Social | 1 | 0 |
| 62 | 3 | ami_18 | Emotional | 5 | Emotional | 1 | 0 |
| 63 | 1 | ami_12 | Behavioural | 8 | Behavioural | 0.75 | 1.0612781<br>2 |
| 63 | 2 | ami_3 | Social | 5 | Social | 1 | 0 |
| 63 | 3 | ami_18 | Emotional | 5 | Emotional | 1 | 0 |
| 64 | 1 | ami_12 | Behavioural | 7 | Behavioural | 0.7142857<br>1 | 1.1488348<br>5 |
| 64 | 2 | ami_4 | Social | 6 | Social | 0.8333333<br>3 | 0.6500224<br>2 |
| 64 | 3 | ami_18 | Emotional | 5 | Emotional | 1 | 0 |
| 65 | 1 | ami_12 | Behavioural | 7 | Behavioural | 0.8571428<br>6 | 0.5916727<br>8 |
| 65 | 2 | ami_4 | Social | 4 | Social | 1 | 0 |
| 65 | 3 | ami_16 | Emotional | 7 | Emotional | 0.7142857<br>1 | 0.8631205<br>7 |
| 66 | 1 | ami_12 | Behavioural | 7 | Behavioural | 0.8571428<br>6 | 0.5916727<br>8 |
| 66 | 2 | ami_4 | Social | 4 | Social | 1 | 0 |
| 66 | 3 | ami_16 | Emotional | 7 | Emotional | 0.7142857<br>1 | 0.8631205<br>7 |
| 67 | 1 | ami_12 | Behavioural | 7 | Behavioural | 0.8571428<br>6 | 0.5916727<br>8 |
| 67 | 2 | ami_4 | Social | 4 | Social | 1 | 0 |
| 67 | 3 | ami_16 | Emotional | 7 | Emotional | 0.7142857<br>1 | 0.8631205<br>7 |
| 68 | 1 | ami_12 | Behavioural | 7 | Behavioural | 0.8571428<br>6 | 0.5916727<br>8 |
| 68 | 2 | ami_4 | Social | 4 | Social | 1 | 0 |
| 68 | 3 | ami_16 | Emotional | 7 | Emotional | 0.7142857<br>1 | 0.8631205<br>7 |
| 69 | 1 | ami_12 | Behavioural | 7 | Behavioural | 0.8571428<br>6 | 0.5916727<br>8 |
| 69 | 2 | ami_4 | Social | 4 | Social | 1 | 0 |
| 69 | 3 | ami_16 | Emotional | 7 | Emotional | 0.7142857<br>1 | 0.8631205<br>7 |
| 70 | 1 | ami_12 | Behavioural | 7 | Behavioural | 0.8571428<br>6 | 0.5916727<br>8 |
| 70 | 2 | ami_4 | Social | 4 | Social | 1 | 0 |
| 70 | 3 | ami_16 | Emotional | 7 | Emotional | 0.7142857<br>1 | 0.8631205<br>7 |
| 71 | 1 | ami_12 | Behavioural | 7 | Behavioural | 0.8571428<br>6 | 0.5916727<br>8 |
| 71 | 2 | ami_4 | Social | 4 | Social | 1 | 0 |
| 71 | 3 | ami_16 | Emotional | 7 | Emotional | 0.7142857<br>1 | 0.8631205<br>7 |
| 72 | 1 | ami_12 | Behavioural | 7 | Behavioural | 0.8571428<br>6 | 0.5916727<br>8 |
| 72 | 2 | ami_4 | Social | 4 | Social | 1 | 0 |
| 72 | 3 | ami_16 | Emotional | 7 | Emotional | 0.7142857<br>1 | 0.8631205<br>7 |
| 73 | 1 | ami_12 | Behavioural | 7 | Behavioural | 0.8571428<br>6 | 0.5916727<br>8 |
| 73 | 2 | ami_4 | Social | 4 | Social | 1 | 0 |
| 73 | 3 | ami_16 | Emotional | 7 | Emotional | 0.7142857<br>1 | 0.8631205<br>7 |
| 74 | 1 | ami_4 | Social | 4 | Social | 1 | 0 |
| 74 | 2 | ami_16 | Emotional | 7 | Emotional | 0.7142857<br>1 | 0.8631205<br>7 |
| 74 | 3 | ami_12 | Behavioural | 7 | Behavioural | 0.8571428<br>6 | 0.5916727<br>8 |
| 75 | 1 | ami_12 | Behavioural | 6 | Behavioural | 1 | 0 |
| 75 | 2 | ami_4 | Social | 5 | Social | 0.8 | 0.7219280<br>9 |
| 75 | 3 | ami_18 | Emotional | 7 | Emotional | 0.7142857<br>1 | 0.8631205<br>7 |
| 76 | 1 | ami_12 | Behavioural | 6 | Behavioural | 1 | 0 |
| 76 | 2 | ami_3 | Social | 4 | Social | 1 | 0 |
| 76 | 3 | ami_18 | Emotional | 8 | Emotional | 0.75 | 0.8112781<br>2 |
| 77 | 1 | ami_12 | Behavioural | 6 | Behavioural | 1 | 0 |
| 77 | 2 | ami_3 | Social | 6 | Social | 1 | 0 |
| 77 | 3 | ami_18 | Emotional | 6 | Emotional | 1 | 0 |
| 78 | 1 | ami_12 | Behavioural | 6 | Behavioural | 1 | 0 |
| 78 | 2 | ami_3 | Social | 6 | Social | 1 | 0 |
| 78 | 3 | ami_18 | Emotional | 6 | Emotional | 1 | 0 |
| 79 | 1 | ami_12 | Behavioural | 7 | Behavioural | 0.8571428<br>6 | 0.5916727<br>8 |
| 79 | 2 | ami_3 | Social | 6 | Social | 1 | 0 |
| 79 | 3 | ami_18 | Emotional | 5 | Emotional | 1 | 0 |
| 80 | 1 | ami_12 | Behavioural | 7 | Behavioural | 0.8571428<br>6 | 0.5916727<br>8 |
| 80 | 2 | ami_3 | Social | 6 | Social | 1 | 0 |
| 80 | 3 | ami_16 | Emotional | 5 | Emotional | 1 | 0 |
| 81 | 1 | ami_12 | Behavioural | 7 | Behavioural | 0.8571428<br>6 | 0.5916727<br>8 |
| 81 | 2 | ami_3 | Social | 6 | Social | 1 | 0 |
| 81 | 3 | ami_16 | Emotional | 5 | Emotional | 1 | 0 |
| 82 | 1 | ami_12 | Behavioural | 7 | Behavioural | 0.8571428<br>6 | 0.5916727<br>8 |
| 82 | 2 | ami_3 | Social | 6 | Social | 1 | 0 |
| 82 | 3 | ami_16 | Emotional | 5 | Emotional | 1 | 0 |
| 83 | 1 | ami_11 | Behavioural | 4 | Behavioural | 0.75 | 0.8112781<br>2 |
| 83 | 2 | ami_3 | Social | 6 | Social | 0.8333333<br>3 | 0.6500224<br>2 |
| 83 | 3 | ami_10 | Behavioural | 2 | Behavioural | 0.5 | 1 |
| 83 | 4 | ami_16 | Emotional | 6 | Emotional | 0.8333333<br>3 | 0.6500224<br>2 |
| 84 | 1 | ami_11 | Behavioural | 3 | Behavioural | 1 | 0 |
| 84 | 2 | ami_10 | Behavioural | 2 | Behavioural | 0.5 | 1 |
| 84 | 3 | ami_3 | Social | 7 | Social | 0.7142857<br>1 | 1.1488348<br>5 |
| 84 | 4 | ami_16 | Emotional | 6 | Emotional | 0.8333333<br>3 | 0.6500224<br>2 |
| 85 | 1 | ami_11 | Behavioural | 3 | Behavioural | 1 | 0 |
| 85 | 2 | ami_10 | Behavioural | 2 | Behavioural | 0.5 | 1 |
| 85 | 3 | ami_3 | Social | 7 | Social | 0.7142857<br>1 | 1.1488348<br>5 |
| 85 | 4 | ami_16 | Emotional | 6 | Emotional | 0.8333333<br>3 | 0.6500224<br>2 |
| 86 | 1 | ami_16 | Emotional | 8 | Emotional | 0.625 | 0.954434 |
| 86 | 2 | ami_14 | Social | 3 | Behavioural | 0.6666666<br>7 | 0.9182958<br>3 |
| 86 | 3 | ami_3 | Social | 7 | Social | 0.7142857<br>1 | 1.1488348<br>5 |
| 87 | 1 | ami_12 | Behavioural | 9 | Emotional | 0.5555555<br>6 | 0.9910760<br>6 |
| 87 | 2 | ami_5 | Behavioural | 2 | Behavioural | 0.5 | 1 |
| 87 | 3 | ami_3 | Social | 7 | Social | 0.7142857<br>1 | 1.1488348<br>5 |
| 88 | 1 | ami_3 | Social | 7 | Social | 0.7142857<br>1 | 1.1488348<br>5 |
| 88 | 2 | ami_12 | Behavioural | 11 | Behavioural | 0.4545454<br>5 | 1.3485879 |
| 89 | 1 | ami_3 | Social | 7 | Social | 0.7142857<br>1 | 1.1488348<br>5 |
| 89 | 2 | ami_12 | Behavioural | 11 | Behavioural | 0.4545454<br>5 | 1.3485879 |

## Footnotes

1 Although the majority of the NCD cohort were older adults, 12 patients were aged between 23 and 38 years. These comprised individuals with subjective cognitive decline (SCD) or autoimmune encephalitis (AIE), conditions that can present at younger ages. We note this here to clarify the presence of younger participants within the NCD group.

2 We included individuals with Subjective Cognitive Decline (SCD) within the NCDs cohort because SCD is widely recognised as part of the neurocognitive disorder spectrum: it carries elevated risk for progression to Mild Cognitive Impairment and dementia, and is often considered an at-risk or prodromal stage in clinical and research frameworks.

